# Vaccines at Velocity: Evaluating Potential Lives Saved by Earlier Vaccination in the COVID-19 Pandemic

**DOI:** 10.1101/2023.06.16.23291442

**Authors:** Witold Więcek, David Johnston, Tomas Dulka, Danny Toomey, Enlli Lewis

## Abstract

Fast development of COVID-19 vaccines likely averted millions of deaths. We estimate how many more lives could have been saved if safe and effective vaccines were available earlier in the pandemic, in particular, before the epidemic waves in winter of 2020. We fit an epidemiological model informed by retrospective data and simulate counterfactual vaccination scenarios for the United Kingdom and the United States in which vaccines are available between 30 and 90 days earlier. We find that up to 1 July 2021 reductions in mortality range from 10,000 to 48,000 in the UK and 53,000 to 130,000 in the US, depending on when vaccinations start. This corresponds to a maximum of 7.1 and 4 deaths averted per 10,000 people in the UK and US respectively, or a reduction in overall deaths of 50% and 32%. We find that our model is sensitive to uncertain vaccine parameters and benefits depend on the time horizon of the analysis. However, the large average reductions we estimate suggests that it is highly cost-effective to make large investments in strategies to expedite vaccine availability.

## 1 Introduction

The development and testing of vaccines for COVID-19 progressed at an unprecedented pace, with vaccinations approved in some countries within less than a year from the start of the pandemic. However, even with accelerated approvals the COVID-19 pandemic had a devastating impact on global health and economies. In the United States and United Kingdom alone, hundreds of thousands of lives were lost in the period it took to approve and complete mass vaccinations.

It has been suggested that various interventions could have led to the vaccines becoming available even earlier. For example, in the United States some have pointed out that while the clinical trials were conducted efficiently, the emergency use authorization process still involved a gap of several weeks between completing phase 3 studies and the start of mass vaccination. Also in the US, others argued that vaccines should have been approved earlier based on the FDA’s “animal rule” and compassionate use pathways for certain groups.^1^ Another proposal to accelerate vaccine availability involved use of human challenge trials (HCTs) to speed up generation of vaccine efficacy results.^2^

Abstracting from the particular way that vaccines might have been made available earlier and feasibility or risks of any of the possible approaches, it is pertinent to ask what the possible health benefits of such an approach would have been. A priori both health and economic benefits seem likely to be significant. For example, economic research into accelerating COVID vaccine availability has suggested that these benefits may be orders of magnitude larger than the costs associated with acceleration (Castillo et al., 2021).

Despite this, there has been relatively little research on the impact of speed on health benefits, particularly in terms of earlier approval and by using epidemiological modelling. A recent review paper found only one such quantitative estimate for mortality (Więcek, 2022). This estimate, from Berry et al. (2020), was based on prospective simulation done early in the pandemic, which understandably overestimated timelines for vaccine field trials and was not able to use information about time-course of the COVID-19 pandemic in 2020 and 2021 to calibrate the model.^3^

In this paper, we aim to provide a simple revision to this approach, where we use retrospective data to quantify the potential health benefits (which we measure in terms of deaths averted) of accelerating the availability of COVID-19 vaccines. More concretely, we proceed in two stages. First, we calibrate an epidemiological model informed by retrospective analysis of data on observed vaccinations and mortality. Then, considering the actual vaccine development and approval timelines and comparing them to counterfactual acceleration scenarios, we simulate counterfactual epidemics and their associated mortality.

It is important to note that we consider hypothetical accelerated timelines, rather than evaluating risk-benefits from implementing concrete innovations which in some cases may trade off risk (less precise information on efficacy or safety) for speed. We are interested only in the magnitude of potential benefits. For this reason, we do not consider health risks to the general population that accelerating approval of less effective or safe vaccines would result in (Chandra et al., 2022).

We calculate the health benefits for two countries only, the United States and the United Kingdom.^4^ They are large developed countries that were at the front of the queue for vaccine deliveries and therefore would have been among those most likely to benefit from accelerated testing. However, there are also considerable differences in timing of waves, circulating variants, speed of vaccination, adoption of boosters, and vaccine prioritization, to name a few. Therefore it’s instructive to compare the two cases. As we will discuss throughout this article, we contrast these two countries to provide an illustrative example, because they differed in the nature of epidemics (timing of waves, different strains), non-pharmaceutical interventions, and vaccination policy and uptake.

Our epidemiological modelling approach simplifies many relevant real-world factors, including individual behaviors and responses to public health interventions. Therefore, we also conduct some sensitivity analyses and discuss some potential unknowns associated with accelerating vaccine approvals, such as (1) less precise information on efficacy or safety, (2) impact on public confidence in vaccines, (3) the potential for production and distribution constraints to nullify the benefits of accelerated testing.

## 2 Vaccinations and mortality under status quo

Figure 1 shows daily vaccinations and deaths attributable to COVID in the US and the UK.^5^ Between the start and the end of 2021 (that is, within roughly a year of mass vaccination campaigns), 470,000 deaths occurred in the US and 75,000 in the UK, more than doubling the death tolls compared to 2020. 200,000 (US) and 52,000 (UK) of these occurred in the first quarter of 2021, as the vaccinations were picking up speed.

**Figure 1:**
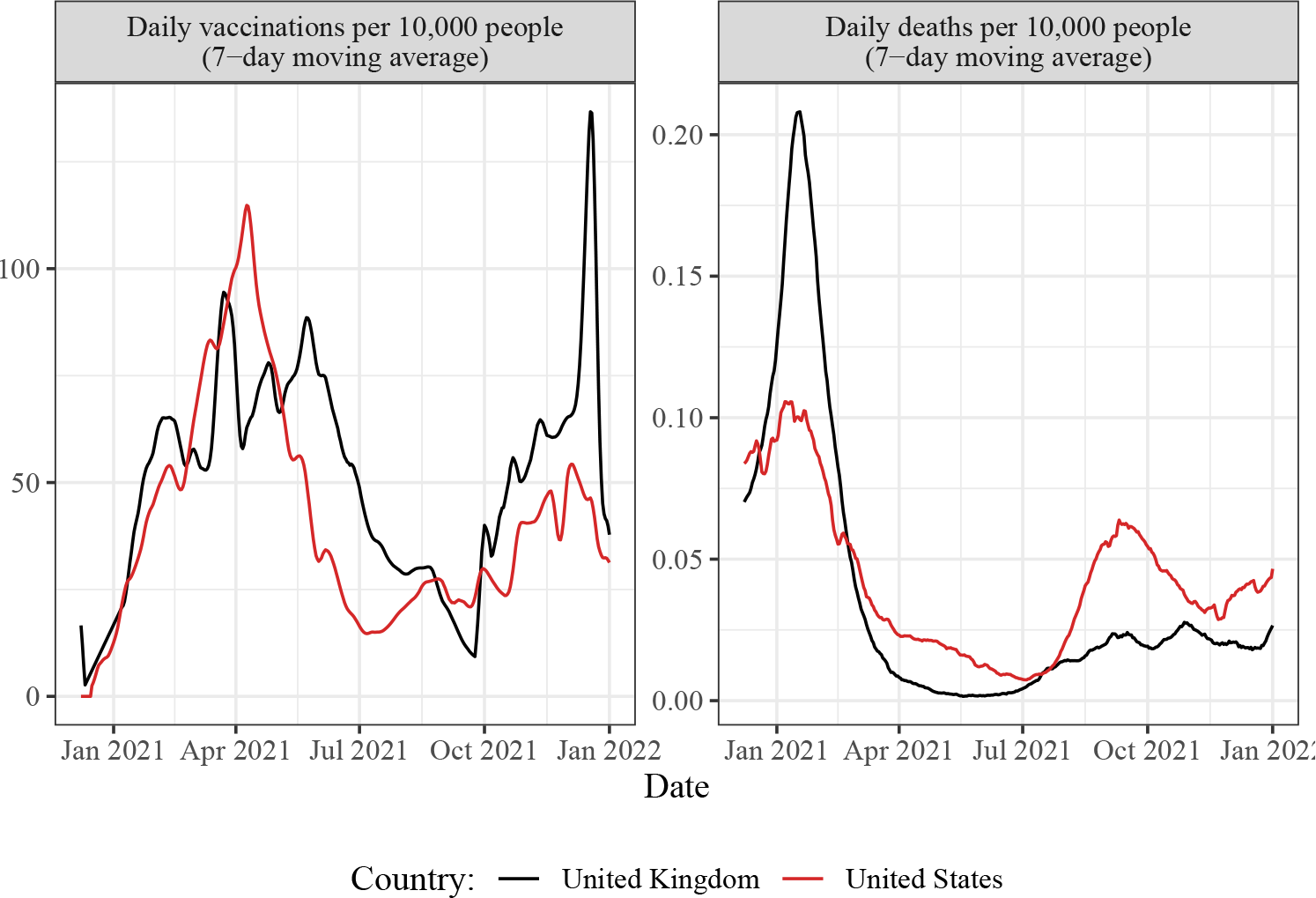
Data on daily vaccinations and deaths in the UK and the US in 2021

Note that severity of epidemic waves was different across these two countries: the UK experienced a very sharp rise in deaths in early 2021 compared to the milder peak in late 2021. This pattern is also present but much less pronounced in the US. In both countries the daily vaccinations only leveled off in early summer of 2021, due to distribution capacity lagging behind demand.

What share of those deaths could have been averted if mass vaccination campaigns started earlier, thanks to earlier approval of the first available vaccines (those manufactured by Pfizer and Moderna in both countries, plus AstraZeneca vaccine in the UK)? One way to address this question is to use counterfactual simulations from epidemiological models.

Watson et al. (2022b) have previously calculated that vaccinations (compared to the hypothetical lack of vaccination) have averted up to 20 million deaths worldwide including up to 2 million deaths in the US and up to 0.5 million deaths in the UK in their first year of use.^6^ According to that calculation, vaccines prevented approximately 2-3 deaths for each death that occurred. In the last three months of 2020, about 150,000 COVID-19-related deaths occurred in the US and about 40,000 in the UK. Therefore a simplistic parallel argument (assuming two lives saved for each death) would suggest that 100,000 lives could have been saved in that period in these two countries alone if vaccines were available.

However, any back-of-envelope calculation on health impacts of accelerating vaccines ignores crucial and obvious epidemiological factors which determine the degree of success of mass vaccination campaigns. First, the reduction in risk is not immediate. It takes time to distribute vaccines and for the vaccinated individuals to develop immunity. Second, even taking the above into account, the impact is not uniform over time due to the risk of infection changing. While for COVID-19 it was not a realistic hope to achieve herd immunity using mass vaccination, epidemiological effects of the vaccine (i.e. breaking transmission) still mean that the impacts are different depending on changes in effective reproductive number of the virus: in other words, whether the vaccinations occur before, during or after a wave of infections. Third, due to gradual waning of immunity and endemic nature of COVID-19, mass vaccination leads to a possibility of a “rebound” in infections and deaths in subsequent waves.^7^

Therefore to address our research question, we use an epidemiological model, one which is a slight modification of the model used by Watson et al. (2022b) which adds support for booster vaccinations and strain-dependent efficacy^8^. However, before turning to the model, we first consider the design of the counterfactual scenarios.

## 3 Vaccine trials and approvals

In building the counterfactual scenarios and epidemiological modelling we focus on vaccines produced by Moderna, Pfizer, and AstraZeneca, which were the main vaccines approved both in the United Kingdom and the United States (with exception of AstraZeneca).

Vaccine development during the pandemic was unusually fast. For the vaccines we discuss it took about nine months from the start of first human trials to vaccine authorizations. Phase II trials began roughly in May 2020.^9^ Following that milestone, to approve a vaccine, safety and efficacy data would be needed. In considering the accelerated approval timelines, we make two considerations: (1) the same safety data could have been generated earlier, by starting large safety trials earlier; and (2) efficacy data could have been generated by HCT or some other means.

In autumn of 2020 the FDA guidance requested that the majority of trial participants need to have at least two months of safety data from the second dose of a vaccine (Herper and Branswell, 2020). Given 3-4 weeks between doses, an optimistic timeline in which safety assessment could be completed without compromising on this rule is late August 2020, roughly three months before Moderna and Pfizer requested emergency use authorization for their vaccines.^10^ This places a potential approval (conditional on proving efficacy) sometime in September.

The second component of vaccine trial length is testing efficacy. While this could have been accelerated in the same way as generating safety data, by investing in larger trials much earlier in the pandemic, it may also be instructive to also consider a hypothetical HCT. With an HCT, testing only takes up to eight weeks (administration of two doses, followed by two weeks’ wait to build up immune response, followed by about two weeks of exposure and measurement of response). In the case of a controlled exposure study with a manufactured challenge agent, the wait would likely be considerably longer and it’s hard to assess whether such an efficacy HCT could generate results faster than a typical field trial. On the other hand, an alternative such as a natural exposure trial (which does not require manufacturing a challenge agent) could have been completed by September 2020.^11 12^

With the above in mind we choose three illustrative counterfactual approvals scenarios, where vaccinations occur 30, 60, or 90 days earlier compared to the actual timeline. These counterfactual approval timelines do not correspond to any particular trial design: our objective here was primarily to crudely identify a realistic range of counterfactual approval dates.

## 4 Hypothesising vaccination rates

We summarize some of the relevant evidence on vaccine demand and production in Appendix A. Obviously approving vaccines faster is only beneficial if they can be distributed in sufficient numbers and people are willing to take them. Therefore, to derive the time-course of vaccinations under our hypothetical vaccine approval timelines, we consider demand, production, and distribution constraints over time.

In all cases we start from the observed vaccination rates (partially and fully vaccinated) based on Our World in Data (Mathieu et al., 2021). The first COVID vaccine was administered outside of a clinical trial on December 14th 2020 in the US and December 8th 2020 in the UK. We then make the following adjustments to create counterfactual scenarios:

### Demand

Early on in the vaccination campaigns, the demand for vaccines far outstripped the supply (see Appendix A for summary of evidence on vaccine hesitancy). However, it is possible that under the hypothetical accelerated approval timelines the demand would have decreased in the short term due to higher vaccine hesitancy. However, once mass vaccination is underway, additional safety and effectiveness data can be derived quickly through a combination of real world data and post-approval phase 4 studies (Dagan et al., 2021). Therefore, it’s best to assume that any possible impact is not on total demand, but on the rate of vaccinations (the rate at which the number of individuals concerned about safety or efficacy decreases). Additionally, the overall vaccine hesitancy in the US and the UK in autumn of 2020 was low and did not vary much over time. In summary, in our base case we assume no differences to demand, but we conduct an additional sensitivity analysis which adjusts vaccination rate downward, by 5% or by 10%.

### Distribution

For the base case we assumed that distribution constraints would be unchanged for each counterfactual.

### Production

We used publicly available information on manufacturing of Pfizer and Moderna vaccines. We assumed a simple linear scale-up of production between March and July 2020 and then constant daily production until the period two weeks before vaccine approvals, in a way that matches the total reported production numbers until end of 2020 (see Appendix A for details and illustration). When we shift the vaccination dates by 30, 60, or 90 days, we do not change production numbers.^13^

Crucially, we assume that available vaccines are distributed to the first country that will make use of them. This means that for the UK the production constraint does not bind, because total manufacturing capacity quickly exceeds the UK population. In the case of the United States we find that the production constraint only binds in the case when the vaccination is shifted by 90 days.

## 5 Vaccination and mortality model

Details of epidemiological model and data are given in Appendix C. Here we only provide a short sketch. As mentioned, our base case model is a slight modification of the model by Watson et al., which is an age-stratified Susceptible-Exposed-Infected-Recovered/Dead (SEIRD) model (Watson et al., 2022b). A model of this type can be considered standard and appropriate for epidemiological modelling and simulations in a large population.

Crucially, the model allows for modelling of variants, distinguishes between number of doses as well as infection-and disease-blocking effects of vaccines, and includes waning of (natural and vaccine-induced) immunity. The model includes vaccine prioritization strategies but mixing of groups is homogeneous across ages. Two types of infections, severe and non-severe, are distinguished. We use age-specific probabilities for infection and death.^14^ Speed of waning of immunity (the rate at which people transition to an appropriate model compartment) depends on the number of doses received. The length of natural immunity is, on average, 250 days.

Because the counterfactual reductions in deaths crucially depend on characteristics of the vaccine, which are uncertain, we follow Watson et al. by drawing 100 sets of parameters of vaccine efficacy and duration of protection. Then, for each set, we calibrate the base case model by fitting it to observed mortality (confirmed cases), derived from the curated dataset created by Dong et al. (2020), and vaccination data from Our World In Data in the period from early 2020 to 1 January 2022 (Mathieu et al., 2021). This provides us with an estimate of the initial number of infections and reproduction number of the virus over time. For each such calibration we then simulate counterfactuals and calculate the averted numbers of deaths under each of them. Therefore for every model-derived quantity we report in this paper, we give both the mean and a 95% interval, calculated over the 100 simulations.

### 5.1 Alternative definition of outcomes

In addition to the base case, we report outcomes of the same model but at different times or fitted to different mortality data:

- **Cut-off dates for analysis:** A major source of variation in the estimates of deaths under accelerated vaccine timelines is the degree of the “rebound effect” which we mentioned earlier. In particular, if we continue the simulation until a winter peak is reached, we sometimes see more deaths under the counterfactual than the baseline. This happens because when people receive vaccines earlier, their effects also wane faster. The size of this rebound has a substantial impact on the overall estimated benefits of accelerated vaccine timelines. Therefore in reporting counterfactual scenarios we calculate health impacts up to 1 July 2021, 1 April 2021, and 1 January 2022.
- **Fitting the model to excess deaths rather than reported deaths:** An epidemiological model which focuses on COVID-19-attributable deaths (our base case) will likely fit better to data, but it may obviously underestimate the total burden of disease. Therefore, following Watson et al. we also report results for an alternative pandemic model which was fit to excess rather than reported deaths.

### 5.2 Sensitivity analyses

We present details of various sensitivity analyses in Appendix D. There are many reasons why the model that we use may not adequately capture the impact of making vaccines available earlier.

First, the SEIRD model is based on an idealization of transmission and vaccination dynamics and there may be features of the true pandemic dynamics that are not adequately captured by this model, but are critical for counterfactual modelling. For example, simplification of modelling of social mixing^15^ or behavioral adjustment in reaction to infection risks may impact the magnitude of counterfactual reductions in mortality.

Moreover, the model depends on many parametric assumptions, and most of these parameters are only identified with substantial uncertainty. Therefore we can ask how varying these parameters may impact the magnitude of health benefits. By doing this, we can understand how misspecifications of the model may affect our conclusions.

We look at the following three parameters, fixing each parameter at the 10th and 90th percentile of their distributions and fitting the mortality model: (1) vaccine efficacy for first two doses, (2) duration of vaccine-induced protection (DVI), (3) duration of naturally acquired immunity (DNI). All three of these parameters are especially important because they impact the number of susceptible individuals over several consecutive epidemic waves. For example, if the natural immunity is longer than we assumed in the model, the rebound effect in subsequent peaks will be less.

We cannot hope to adequately capture all sources of uncertainty present in our model. However, if there was large variation in modeled deaths averted when choosing between similarly sensible assumptions, this should increase our skepticism in model conclusions.

Lastly, we consider additional sensitivity analysis assuming decreased vaccination demand due to vaccine hesitancy of 5% or 10% under counterfactual scenarios.

## 6 Estimated mortality reductions

Numbers of vaccine doses distributed under the base case and counterfactual models are given in Table 1 and Figure 2. Focusing on our chosen period of up to 1 July 2021, we find 9% increase in the US in the doses administered and corresponding 20% increase for the UK, under the most optimistic scenario of vaccines being available 90 days earlier. However, the differences are much larger in the earlier periods. For example, up to 1 April 2021, the corresponding increases are 68% and 105% for the US and UK respectively.

**Figure 2:**
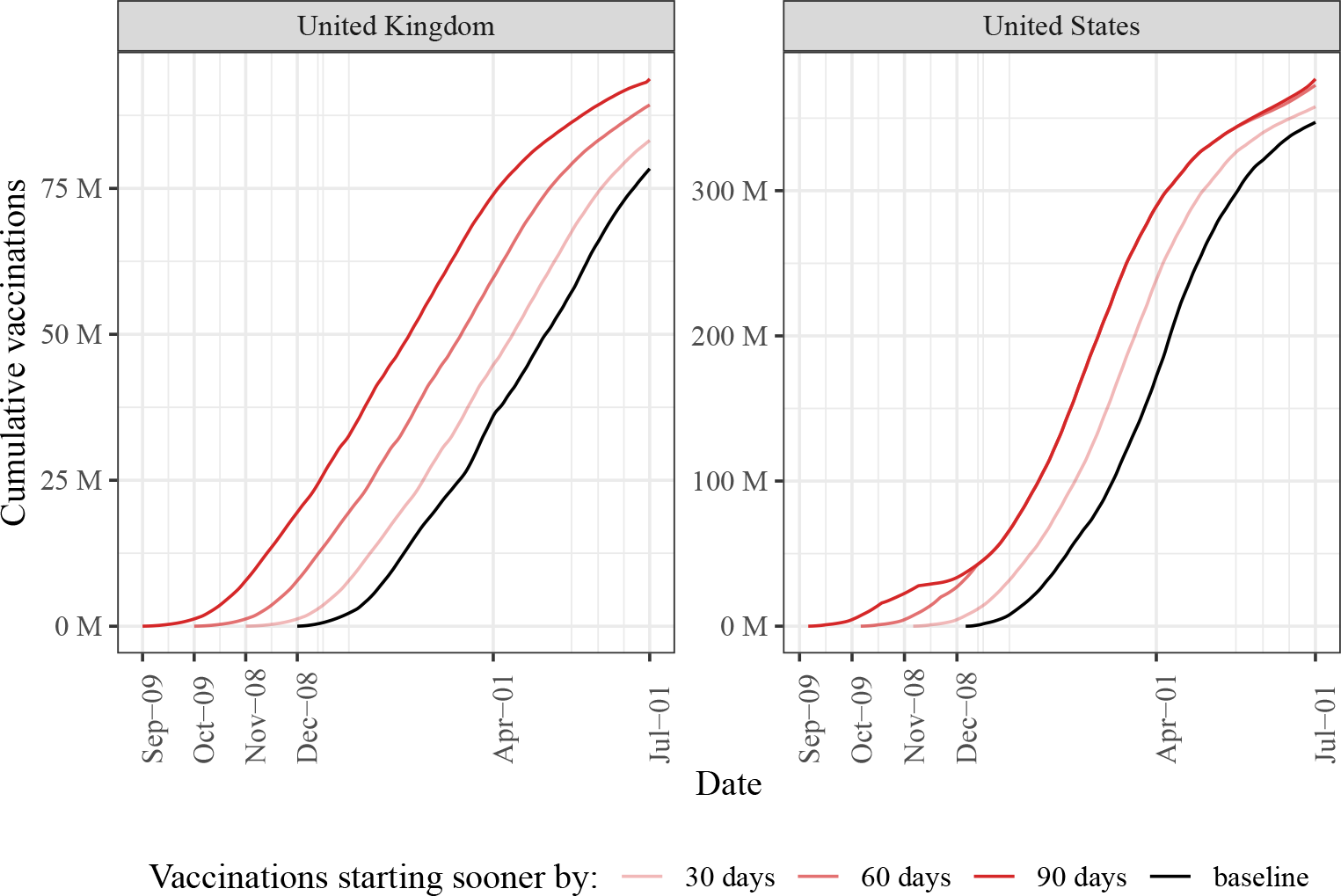
Cumulative vaccinations under baseline and counterfactual scenarios

**Table 1:** Total vaccinations under baseline and counterfactual scenarios

| Counterfactual scenario | 2021-01-01 | 2021-04-01 | 2021-07-01 |
| --- | --- | --- | --- |
| **United Kingdom** |  |  |  |
| Baseline | 1,462,757 | 36,015,897 | 78,366,737 |
| Vaccines 30 days sooner | 5,868,579 | 44,767,107 | 83,205,069 |
| Vaccines 60 days sooner | 17,132,520 | 59,668,154 | 89,288,187 |
| Vaccines 90 days sooner | 30,302,416 | 73,937,217 | 93,731,611 |
| **United States** |  |  |  |
| Baseline | 4,521,988 | 172,124,225 | 347,123,032 |
| Vaccines 30 days sooner | 23,889,241 | 238,670,399 | 357,876,350 |
| Vaccines 60 days sooner | 56,107,462 | 289,245,438 | 372,702,522 |
| Vaccines 90 days sooner | 56,107,489 | 289,245,510 | 376,955,754 |

Model fit is presented in Figure 2 and comparison of counterfactual scenarios over time in Figure 3. We summarize cumulative reductions in mortality in Table 2. In our main period of interest, up to 1 July 2021, our model suggests that for the UK accelerating availability of vaccines would have averted between 10,000 (30-day acceleration; lower 95% bound is 6,000) and 48,000 (90-day acceleration; upper 95% bound is 67,000) deaths (Figure S7, Figure S6). This translates to 1.5 and 7.1 deaths averted per 10,000 individuals or a reduction in overall deaths by 21% to 54%% in the period between start of vaccinations and 1 July 2021. As a reminder, lower and upper bounds correspond to uncertainty in vaccine parameters.

**Figure 3:**
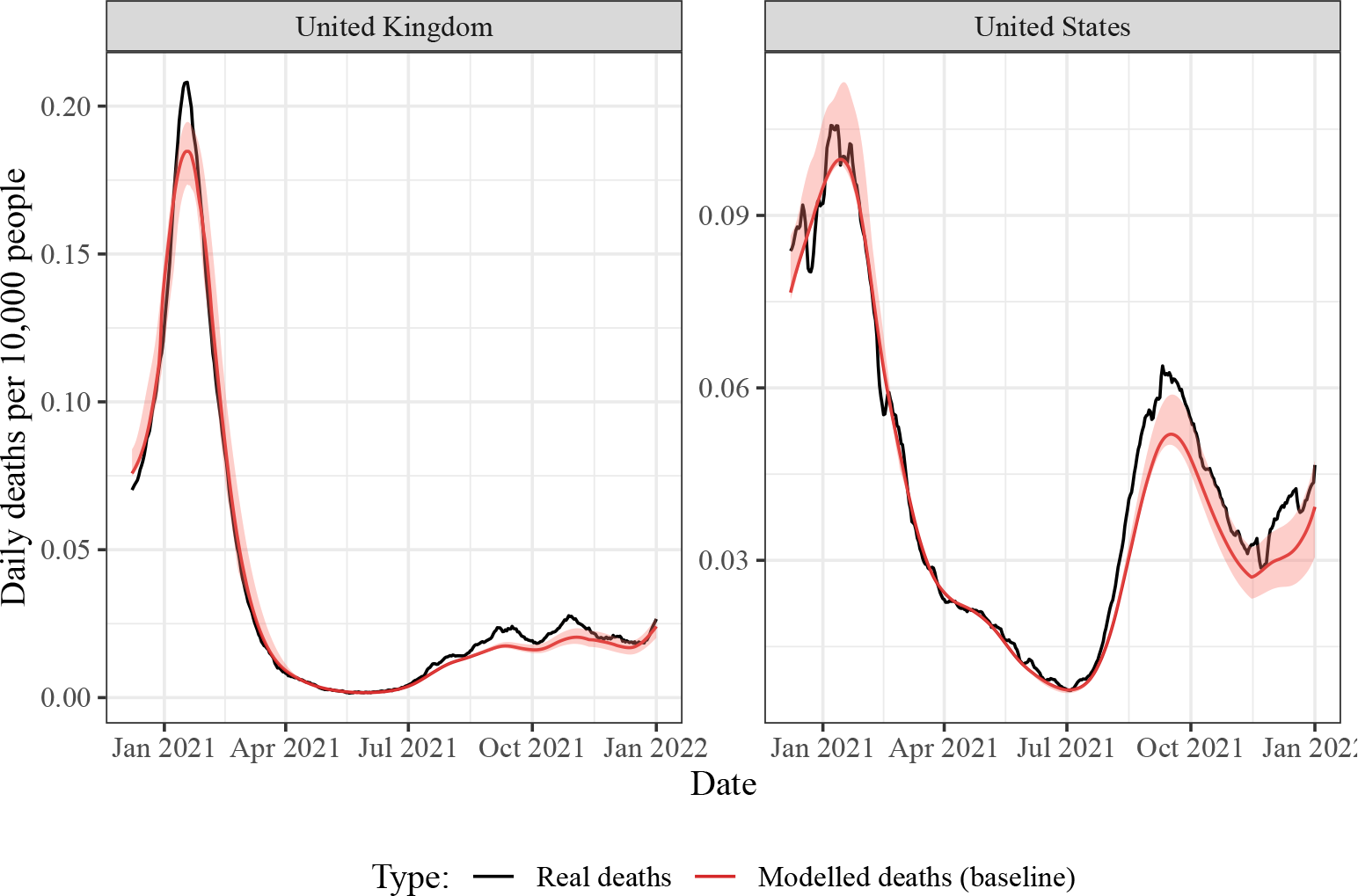
Comparison of real and modelled (baseline) deaths over time. Shaded areas indicate 95% confidence interval.

**Table 2:** Total averted deaths under counterfactual scenarios, based on reported deaths. 95% intervals are calculated over 100 simulation runs which vary vaccine parameters and duration of natural immunity (see Table S1 in Appendix for details). More details are given in Table S7.

| Counterfactual scenario | Deaths averted | Per 10,000 people |
| --- | --- | --- |
| **United Kingdom to April 2021** |  |  |
| 30 days sooner | 10,728 [6,627; 14,991] | 1.60 [0.99; 2.23] |
| 60 days sooner | 29,922 [21,040; 43,472] | 4.46 [3.14; 6.48] |
| 90 days sooner | 46,665 [37,966; 67,151] | 6.96 [5.66; 10.01] |
| **United Kingdom to July 2021** |  |  |
| 30 days sooner | 9,952 [6,026; 13,949] | 1.48 [0.90; 2.08] |
| 60 days sooner | 30,516 [21,967; 43,452] | 4.55 [3.27; 6.48] |
| 90 days sooner | 47,953 [39,529; 67,086] | 7.15 [5.89; 10.00] |
| **United Kingdom to Jan 2022** |  |  |
| 30 days sooner | 8,019 [-2,350; 17,181] | 1.20 [-0.35; 2.56] |
| 60 days sooner | 41,389 [35,940; 49,167] | 6.17 [5.36; 7.33] |
| 90 days sooner | 65,485 [60,730; 77,178] | 9.76 [9.05; 11.51] |
| **United States to April 2021** |  |  |
| 30 days sooner | 35,207 [25,342; 63,091] | 1.06 [0.76; 1.90] |
| 60 days sooner | 87,601 [67,671; 143,053] | 2.64 [2.04; 4.32] |
| 90 days sooner | 107,134 [86,198; 167,087] | 3.23 [2.60; 5.04] |
| **United States to July 2021** |  |  |
| 30 days sooner | 53,342 [45,493; 73,576] | 1.61 [1.37; 2.22] |
| 60 days sooner | 115,895 [99,725; 152,977] | 3.50 [3.01; 4.61] |
| 90 days sooner | 132,919 [117,226; 173,179] | 4.01 [3.54; 5.22] |
| **United States to Jan 2022** |  |  |
| 30 days sooner | 45,196 [3,320; 101,695] | 1.36 [0.10; 3.07] |
| 60 days sooner | 120,655 [-32,178; 221,714] | 3.64 [-0.97; 6.69] |
| 90 days sooner | 175,230 [56,961; 255,219] | 5.29 [1.72; 7.70] |

In the US, corresponding numbers of deaths averted are 53,000 (30-day acceleration; lower 95% bound of 45,000) and 130,000 (90-day acceleration; upper 95% bound of 170,000) for 30-day and 90-day acceleration respectively. This gives a range of 1.6-7.1 deaths averted per 10,000 or a reduction in overall deaths by 15% to 32% in the period between start of vaccinations and 1 July 2021.

As these numbers suggest, the simulations for the UK are typically more sensitive to how much earlier the vaccines become available, likely due to higher vaccine uptake in the UK than in the US (also uptake of boosters: about 50% in the UK compared to only about 25% in the US up to 1 January 2022) and no binding constraints on vaccine availability in the UK even under the most optimistic scenario, unlike in the US.

On the other hand, reductions in the UK are generally lower than in the US. This is likely due to the fact that benefits of vaccination are biggest if mass vaccination precedes rise in cases (for illustration see Fig S6 in Więcek et al. (2022)). In the UK numbers of cases were already rapidly rising in winter 2020 and at the lower end of speed-up counterfactuals (30 days) only a small proportion of cases would be averted, because a small proportion of population would have been vaccinated before epidemic peak (see Figure 4).

**Figure 4:**
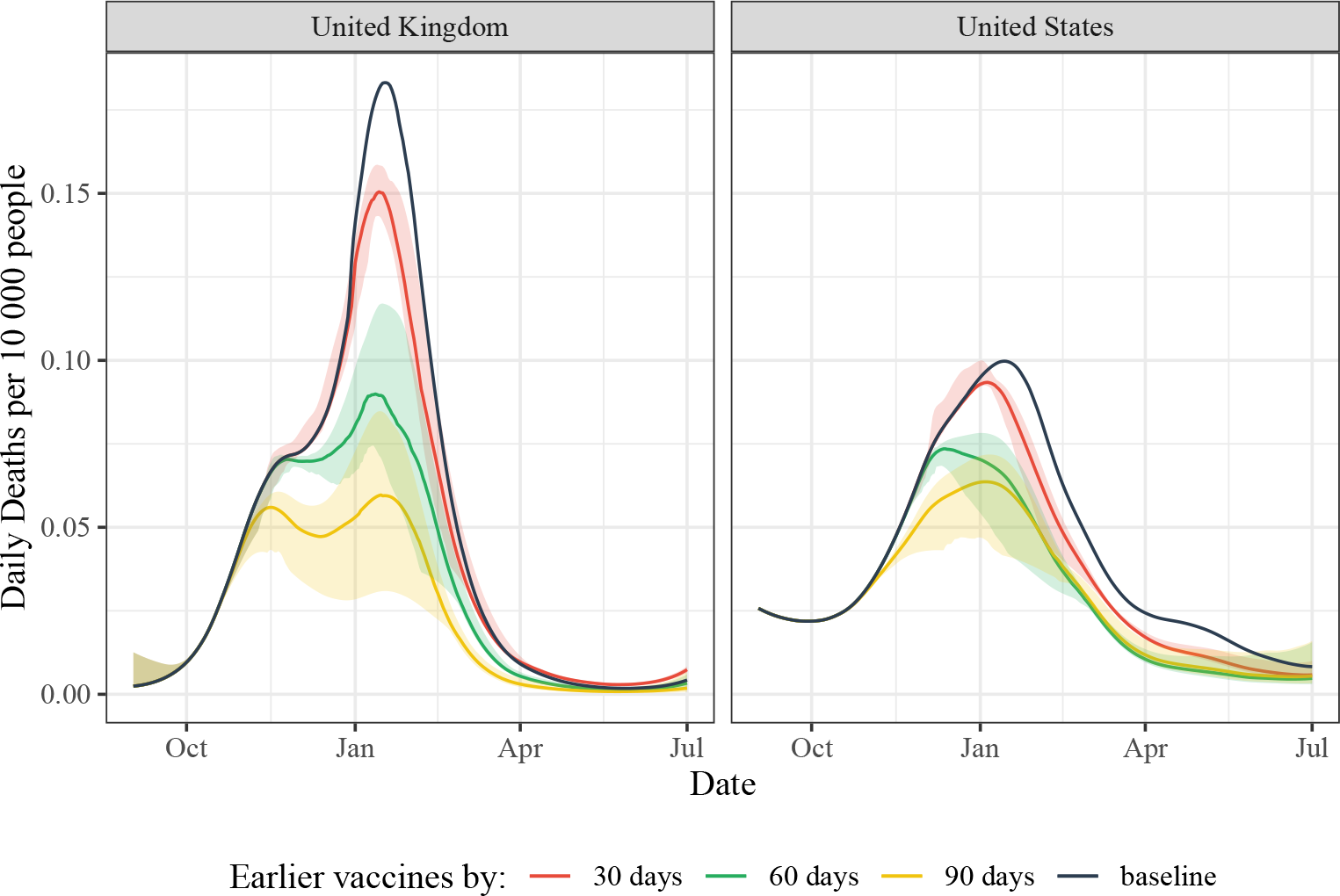
Daily deaths under baseline and counterfactual scenarios. Shaded area indicates 95% confidence interval. Cumulative reductions are shown in the Appendix.

We also noted that while the simulations in the US are affected by the acceleration factor a bit less, within each counterfactual they are more uncertain. This is possibly also explained by the timing of epidemic peaks. Whereas in the UK the crucial parameter is timing of mass vaccination compared to epidemic peak, in the US the deaths are spread more evenly over time and therefore more sensitive to vaccine parameters such as efficacy or duration of protection.

### 6.1 Results of alternative model specifications

Here we summarize our main findings.

#### Cut-off dates for calculation of deaths averted

If we extend the modeled period up to 1 January 2022, the average number of deaths averted generally increases (8,000 to 65,000 for the UK, 45,000 to 65,000 for the US; the ranges represent 30-day to 90-day accelerations). However, uncertainty increases too and we see a considerable “rebound” effect, where the number of deaths can even increase under some trajectories when compared to the status quo. This is because the model is very sensitive to duration of protection.

#### Using excess deaths

See Table S11. If using the excess deaths the effects show similar trends, but are generally smaller, sometimes by as much as a third. This diminished effect is somewhat to be expected, since excess deaths measure also includes non-COVID-attributable deaths and may therefore be less sensitive to increasing vaccination.^16^

#### Additional counterfactual analysis considering booster uptake

Noting differences between the UK and the US (also in the extent variability in results), which had widely different levels of uptake of boosters in the considered period, we conducted an additional analysis, where we increased the uptake of third doses in the US to the same level as in the UK. The results are in Table S10. However, we found that this modification does not account for the differences between the countries.

#### Decreased vaccination demand due to vaccine hesitancy

See Table S8 and Table S9. If demand for vaccines was lower under the counterfactual scenario, the mortality reductions still persist, although they are diminished at the lower end of acceleration (30 days counterfactual): with 10% lower demand we estimate 7,600-45,000 deaths averted in the UK (compared to 10,000-48,000 range holding demand constant) up to 1 July 2021 and 40,000-120,000 in the US (compared to 53,000-130,000 range), with the range corresponding to three counterfactuals (30, 60, 90 days). However, if the decreased demand continued past summer, the mortality reductions would be much smaller.

#### Sensitivity to mis-specified vaccine parameters

See Appendix D for details. Typically mis-specifying vaccine efficacy (first two doses), DVI, or DNI led to decrease in estimated benefits, sometimes by a considerable margin (one-third or even half of the reductions seen under the base case), but in a few cases we also saw increases in benefits. Overall these results suggest large sensitivity to parameter misspecification.

## 7 Discussion

Overall, using an epidemiological model we find large health benefits from accelerating availability of vaccines, both in the UK and the US. In our main period of interest, from the start of vaccination up to 1 July 2021, these reductions range on average from 1.5 up to 7.1 lives saved per 10,000 people, depending on country and amount of acceleration of vaccination schedule (30-90 days). These are obviously large numbers, corresponding to 63,000-180,000 lives saved in two countries alone in the modeled period.^17^

However, our results are uncertain and sensitive to choice of the model, as is typical in all such epidemiological modelling studies. Each counterfactual calculation of deaths averted (see Table 2) varied highly, depending on vaccine parameters. Moreover, there are several sources of uncertainty not captured by the 95% intervals we cite. First, as our sensitivity analyses show, model misspecification would have considerable impact on the estimates of deaths averted. Second, the benefits are sensitive to the choice of whether we model reductions in excess deaths or confirmed deaths. Third, relative reductions in mortality depend on the time horizon of the analysis, since in some cases reductions in mortality from the initial wave are balanced by increase in cases in later waves.

In sum, while our results should be seen as indicative of large potential gains, they should not be treated as precise estimates. However, the variability in results underscores how benefits of mass vaccination are highly sensitive to their timing before epidemic waves and the crucial role of sustaining protection during subsequent waves, to avoid a rebound in cases. The difficulty of estimating potential mortality reductions related to the fact that duration of vaccine-induced and natural protection (which drive the rebound) are hard to estimate even retrospectively.

We are not aware of retrospective epidemiological analyses of how mortality changes with earlier start of mass vaccinations, but our overall conclusion of possible large gains from accelerating availability of vaccines fits into a growing literature on benefits of accelerating vaccination. For example, Bjoerkheim and Tabarrok (2022) estimate 40,000 lives in the US could have been saved by making vaccines available 3 months earlier to care home residents. However, their model is not epidemiological and restricted to a particular population. A recent modelling work by Imai et al. (2023), also using retrospective data, suggests that extending interval between doses in the UK (from a 3-week gap to 12 weeks) averted 10,000 deaths in the period up to September 2021, an about 16% decrease in mortality. Many simulations (see, for example, Bubar et al. (2021)) predict large differences in benefits depending on prioritization strategies when vaccine supply is constrained, especially with regards to age. Another epidemiological simulation suggests that 22-47% of deaths could have been averted in COVID waves if the speed of vaccinations was doubled, with significant gains possible even if a less effective vaccine was used, for example via fractional dosing (Więcek et al., 2022).

### 7.1 Do benefits justify investment?

Does the large health benefit found by our model justify investing more into additional trials? Consider the following illustrative example. Firstly, let’s assume that going into Phase II COVID vaccine developers predicted a 33% chance of success.^18^ Secondly, let’s assume some hypothetical innovation (faster field trials, use of HCTs, streamlined approval process) has a 6% chance of accelerating approval by 30 days, 3% by 60 days or 1% by 90 days. In expectation this gives 3,300 lives saved in two countries. If implementing this innovation for all leading vaccine candidates cost 100 million USD total, that’s still about 300 USD per life saved. This figure ignores other health and economic benefits and the fact that a vaccine is a global public good. If we were to factor these in, the benefits would have likely been even orders of magnitude larger.

The 100 million cost quoted here is just a rough estimate. Costs will of course depend on the type of innovation used to accelerate the trial. In the UK, the world’s first coronavirus human challenge study, conducted by hVIVO, which involved up to 90 volunteers, was backed by a £33.6 million UK government investment.^19^ In a pandemic setting these costs will be likely higher, e.g. overall funding for vaccine R&D in the US provided by the government amounted to $1.7 billion, but only part of it was spent on clinical trials (Lalani et al., 2023). Moreover, the probabilities of acceleration that we assumed in the above example (6%, 3%, and 1%) are purely hypothetical.

We focus on two illustrative cases, of the UK and the US. Implicit in our approach is the assumption of the first mover advantage, i.e. that the benefits of speed accrue most in countries that can accelerate regulatory approval the most. For that reason, countries with smaller populations also stand to gain more. For larger countries it’s more likely that the manufacturing constraints will bind them, preventing them from reaping all benefits of accelerating vaccine approvals early on.

### 7.2 Considerations for HCTs of vaccine efficacy

Many of the debates about speeding up vaccine testing have focused on using HCTs to determine efficacy. While our results suggest large benefits of even those clinical trial innovations that have relatively small probability to accelerate approval, it is important to note that there is currently no clear regulatory framework for the use of human challenge trials for vaccine evaluation or licensure (see Baylor (2021)). Establishing a regulatory framework in advance of a pandemic would be key to capitalizing on any possible benefits of human challenge trials, in particular, building consensus on how human challenge trials can be used for facilitating emergency use authorization for a vaccine.

As with any clinical trial, human challenge studies are not without risks, and require a number of ethical considerations. This is especially the case when conducting challenge studies with pandemic pathogens, where the administered pathogen is especially virulent or deadly, without available therapeutics. As such ensuring robust safety measures are in place to protect patient safety is paramount.^20^ These might include robust informed consent processes, ethical assessments by external review boards, and nuanced risk-benefit analyses. Efforts have been underway to clarify the ethics of using human challenges for novel pathogens and pathogens with pandemic potential, resulting in a framework for deciding when it is suitable to conduct human challenge trials and the necessary steps to ensure their ethical conduct (see Williams et al. (2023)). While ethical considerations are important, the direct health risks to trial participants are likely low (see e.g. Manheim et al. (2021)).

## 8 Conclusion

In conclusion, our epidemiological modelling suggests that accelerating COVID-19 vaccine approvals could result in significant health benefits, with potentially thousands of lives saved in just a few months. There are large uncertainties in the model, associated costs, and practical feasibility of any such innovations, especially human challenge trials. However, the potential benefits of faster vaccine availability are very large, especially considering the global public good nature of vaccines.

## Data Availability

All data produced in the present work are available upon reasonable request to the authors.

## Appendices

### A Manufacturing constraints

If vaccines were approved earlier, production capacity would probably have been a major constraint on the number of doses delivered at the beginning of the mass vaccination campaigns.

We make three major assumptions:

- As mentioned in the main text, hypothesizing the extent to which production is a binding constraint is complicated by the fact that new manufacturing capacity (or expansion of existing capacity) was put in place in anticipation of regulatory approval. Therefore it is likely that an earlier approval would have led the vaccine makers to expand production faster. In our base case model we make a conservative assumption of no expansion of capacity under earlier approval.
- While we model the United States and the United Kingdom using separate epidemiological models, we do not differentiate between countries when it comes to manufacturing capacity, reasoning that if a country was able to approve a vaccine earlier it would jump to the front of the vaccine deliveries queue.
- We also ignore AstraZeneca’s vaccine (not available in the US in the modelled period) since the doses manufactured by Pfizer and Moderna vaccines are enough to satisfy demand in the UK in the initial phase of vaccinations. However, as a conservative assumption, we still assume that vaccine effectiveness in the UK is a mix of mRNA vaccines and the one manufactured by AstraZeneca.

With these assumptions in place, we hypothesize the daily vaccine production over 2020. We use information about the actual manufacturing timelines and total vaccine production in the modelled period as follows:

- First, we assume that in 2020 Moderna produced 20 million doses and Pfizer/BioNTech produced 50 million (NPR, 2020, Pfizer (2020b)). Additionally, we know Moderna was producing 500,000 doses a day at the time their vaccine was authorized and assume Pfizer/BioNTech was producing 1 million doses a day at the time they received authorization [NPR (2020)]^21^.
- We assume a linear scale-up of production from March to July, followed by constant production until two weeks before authorization by the FDA. Specifically, we assume Pfizer/BioNTech started working on scale up from their announced partnership on March 17th and Moderna from March 23rd (Pfizer, 2020a, Time (2020), Moderna (2020a)). The start of mass production in July (for calculations we use July 31st) was assumed based on reports that Pfizer might have been ready to mass produce by then, with Moderna entering a partnership with Catalent in June to do their fill-and-finish processing with an estimated start at the beginning of Q3 (Pfizer, Moderna (2020b)).
- Time of vaccine authorization was December 11th Pfizer/BioNTech, and December 18th for Moderna meaning another scale up in manufacturing started on November 27th for Pfizer and December 4th for Moderna (FDA, 2020a, FDA (2020b)). We assume that at that point both companies already knew that they would most likely get authorization and were able to do another linear scale-up to a production level they would maintain until the end of the year.
- After the end of 2020 we assume production is unconstrained. This assumption is not important since even with a 90-day shift of vaccination series the production constraint stops binding before the end of 2020.

Figure S1 shows the hypothetical daily production compared to vaccinations under different acceleration scenarios (status quo or 30 days, 60 days, 90 days faster). In the case of the United Kingdom we find that in all scenarios of earlier vaccination the production constraint does not bind at all, since the amount of manufactured doses is very large compared with their population size. In the case of the United States the production constraints binds, but only in the case when the vaccination is shifted by 90 days.

**Figure S1:**
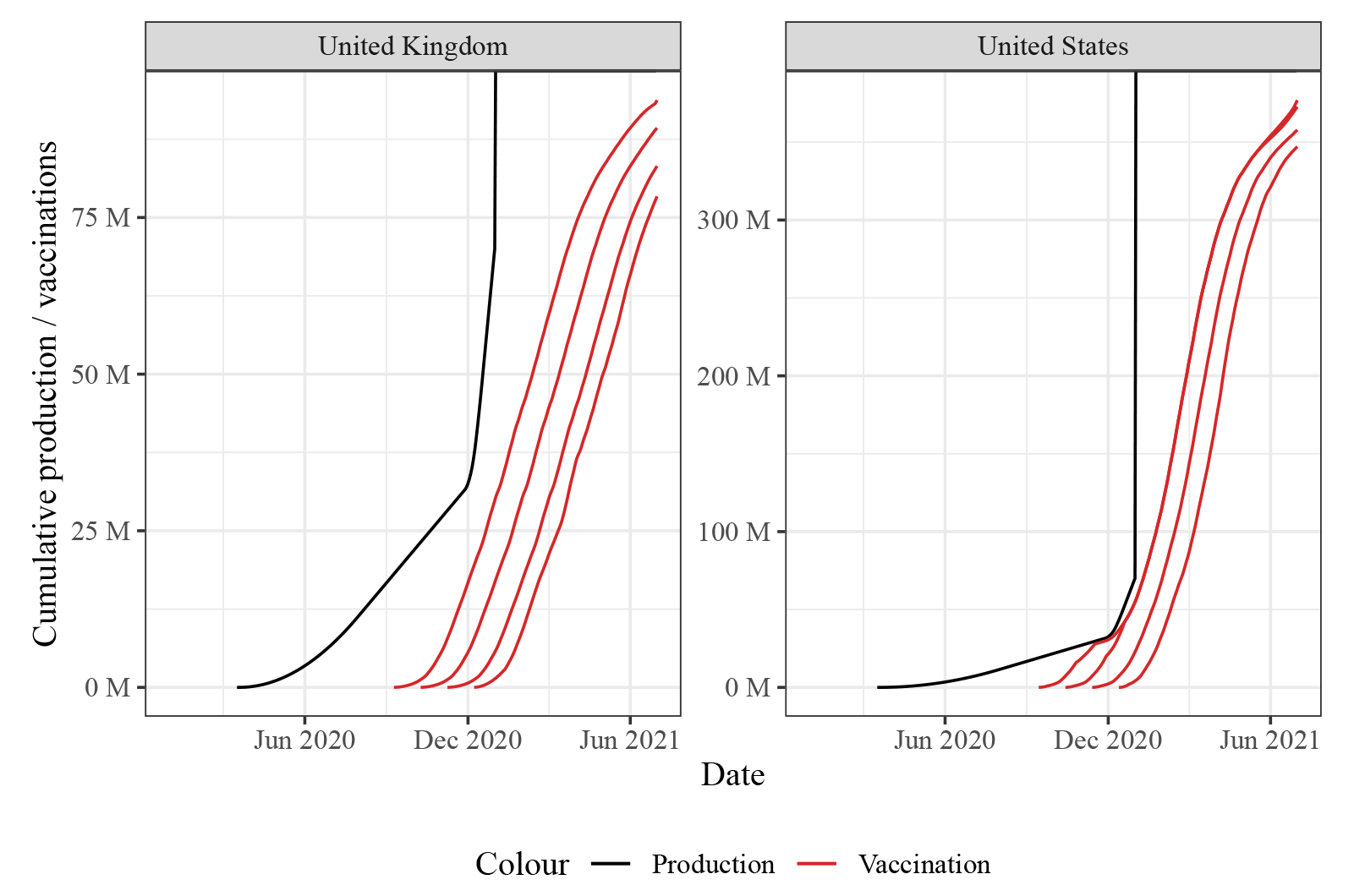
Cumulative vaccine production and distribution in the UK and the US under counterfactual and baseline scenarios. Four lines in red are different acceleration factors, from baseline up to 90 days earlier approval.

### B Vaccine hesitancy

A useful framework for vaccine hesitancy is provided by the “5C’s” model, which considers the following drivers: confidence in vaccine safety, complacency, convenience, risk calculation, and sense of collective responsibility (Machingaidze and Wiysonge, 2021). The factor most clearly affected by a hypothetical earlier approval is confidence in safety (and efficacy). Therefore we used surveys evaluating intention to be vaccinated and confidence in health authorities to approximate the effect of vaccine hesitancy within our time-frame of interest.

In the US and UK, the rate of respondents who intended to be vaccinated when vaccines were approved was 41% and 73% respectively. These proportions at 90 days prior to approval were 39% in the US and 71% in the UK, with the lowest rates of intention to be vaccinated at 36% in October 2020 in the US and 61% in November 2020 in the UK (YouGov, 2023a, Freeman et al. (2022)).

In a separate series of surveys in the US, 85% of respondents had “a lot” or “a fair” amount of confidence in health authorities at time of vaccine approval, compared to 89% in April 2020 (Kennedy, 2022). In the UK, surveys asking respondents their confidence in the country’s health system indicated 73% were confident at time of approval and 77% were confident 90 days prior (YouGov, 2023b). By these estimations, vaccine hesitancy and confidence in health authorities hovered around similar levels during our period of interest.

Dzieciolowska et al. (2021) conducted a study of 2,761 health care workers in Canada in December 2020 to assess vaccine hesitancy, with 19.1% of participants indicating they were not willing to receive the COVID vaccine. Among participants who refused the vaccine, 82% indicated the novelty of the vaccine was very or somewhat important in their decision and 60% indicated that they did not have enough time to make a decision. An approval of vaccines using accelerated timelines could obviously negatively impact hesitancy rooted in vaccine novelty and feelings of needing more time to decide. However, the main hypothetical we discuss in our paper is accelerating efficacy testing and starting safety testing early. This also suggests that while the impact may be non-negligible, it may not be large.

Joshi et al. (2021) found that the global average vaccine acceptance rate was 86% in March 2020, dropped to 54% in July 2020, and increased to 72% in September 2020. By these estimates, if vaccines were approved during summer of 2020, hesitancy may have appreciably impacted uptake. However, it appears that vaccine hesitancy would likely not have impacted uptake if vaccines had been available up to 90 days earlier in the UK and US, with surveys evaluating intention to be vaccinated and trust in health authorities varying by no more than a few percentage points during this time. It is possible these trends would have been impacted by a concern among the general public for vaccines approved quickly, but these effects are difficult to quantify.

### C Epidemiological methods

As discussed in the main text, to model alternative vaccination timelines, we took an existing compartmental model that simulated the COVID-19 pandemic through 2020 and 2021 including the impact of vaccinations (which throughout the paper we refer to as the baseline or base case model) and re-ran the simulation freezing all parameters except the vaccination timelines (the counterfactual models). Here we provide more details on the structure and assumptions of the epidemiological model, as well as its parameters.

The baseline pandemic model builds on earlier work (Watson et al., 2022a). The model is implemented in open source R packages Squire and SirCOVID (Hogan et al., 2021, Walker et al. (2020),Baguelin et al. (2022)). An older version of this model is described in detail in the supplementary material of Walker et al. (2020), and details of many parameters (again, for an older version of the model) can be found in the supplementary material of Watson et al. (2022b). Additional (incomplete) information about the parametrization of the version of the model we used can be found at COVID-19 LMIC Reports documentation (Watson et al., 2022a) and (more up-to-date but less complete) model structure can be found in the Nimue documentation (Winskill et al., 2022). Here we will describe the model in broad terms, without providing all of these details, for which the reader can refer to the original publications.

The epidemiological model is an age stratified SEIRD (susceptible-exposed-infectious-recovered-dead) model. In addition to classifying the population by the disease (SEIRD) status, the model also classifies people by vaccination status: they may be unvaccinated, vaccinated with 1 dose, vaccinated with 2 doses, vaccinated with 2 doses with waned protection, vaccinated 3 doses, or vaccinated with 3 doses with waned protection. People are vaccinated according to vaccination dosage data from Our World in Data (Mathieu et al., 2020) combined with a default model of vaccine prioritization. They progress to “waned” classes according to an assumed rate of waning of vaccine-derived immunity, which is itself time-and dose-number-dependent. The model accounts for separate modes of vaccine action (infection blocking vs disease blocking), variant-dependent efficacy of vaccine and naturally derived immunity and age-dependent vaccination strategies.

While disease progression risks, mortality risks, and vaccination patterns are heterogeneous with age, the model assumes homogeneous mixing of the population. Two kinds of infection are modelled, “severe” and “non-severe”, each with different, age-dependent probabilities of progressing to recovery or death. Individuals who have recovered are assumed to be fully protected by natural immunity for an exponentially random duration which is, on average, around 250 days. Individuals who have been vaccinated have different levels of protection at different points in time depending on the dominant strain, and unless they receive additional doses then protection is considered to have waned after an exponentially random period of time (on average, around 150 days). Vaccinated individuals are also considered to have a reduced likelihood of onward transmission, which also fades when the vaccine protection wanes.

The model features a mix of deterministic parameters and random parameters. We run an ensemble of simulations for each country, sampling the random parameters at the start of each run from pre-defined distributions. The initial number of cases seeding the pandemic in the country and the time series of reproduction numbers of infections in a fully susceptible population *R_t_* is then fit to the observed course of deaths during the time period associated with the simulation.

The distributions of random parameters is given in Table S1. Parameters are sampled from each distribution independently except for the duration of vaccine-derived immunity, which is dependent on vaccine efficacy. 100 sets of parameters are sampled, and epidemic trajectories simulated for each. 95% intervals in our results refer to percentiles among the set of 100 sampled trajectories, so a 95% interval for deaths averted is the pair of numbers given by the 2.5th and the 97.5th percentile of deaths averted among all sampled trajectories. Note that the distributions given here are not exact representations of the models sampling distributions, which involve additional truncation steps.

The model of Watson et al. (2022a) also samples probabilities of hospitalization and mechanical ventilation for each trajectory, but these play no role in our analysis not already captured by the infection fatality rate, and so we haven’t reported on the relevant parameters here.

**Table S1:** Table S1: Distributions from which parameters are sampled. The parameters 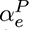, 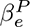 are shape parameters for vaccine platform *P* with associated efficacy *e* (see text for additional details). The parameters *α_d_, β_d_* are shape and rate parameters for distribution of vaccine durations, determined by fitting an antibody decay curve to vaccine efficacies. rescale(*·, f_min_, f_med_, f_max_*) is a function that maps 0 to *f_min_*, 0.5 to *f_med_* and 1 to *f_max_*, linearly interpolating between each. *f_min_, f_med_, f_max_* are respectively the minimum, median, and maximum estimates of age-adjusted infection fatality.

| Parameter | Distribution |
| --- | --- |
| Vaccine efficacy, $V$ | $V \sim \text{Beta}(\alpha_e^P, \beta_e^P), P = \text{vaccine platform})$ |
| Vaccine immunity duration, $D$ | $\frac{1}{\text{Gamma}(\alpha_d, \beta_d)}$ |
| Infection fatality rate with vaccination, $F$ | $F = \text{rescale}(X, f_{\min}, f_{\text{med}}, f_{\max}), X \sim \text{Beta}(2, 2)$ |
| Natural immunity duration, $R$ | $R = \text{Gamma}(20, 4/73)$ |

**Table S2:**
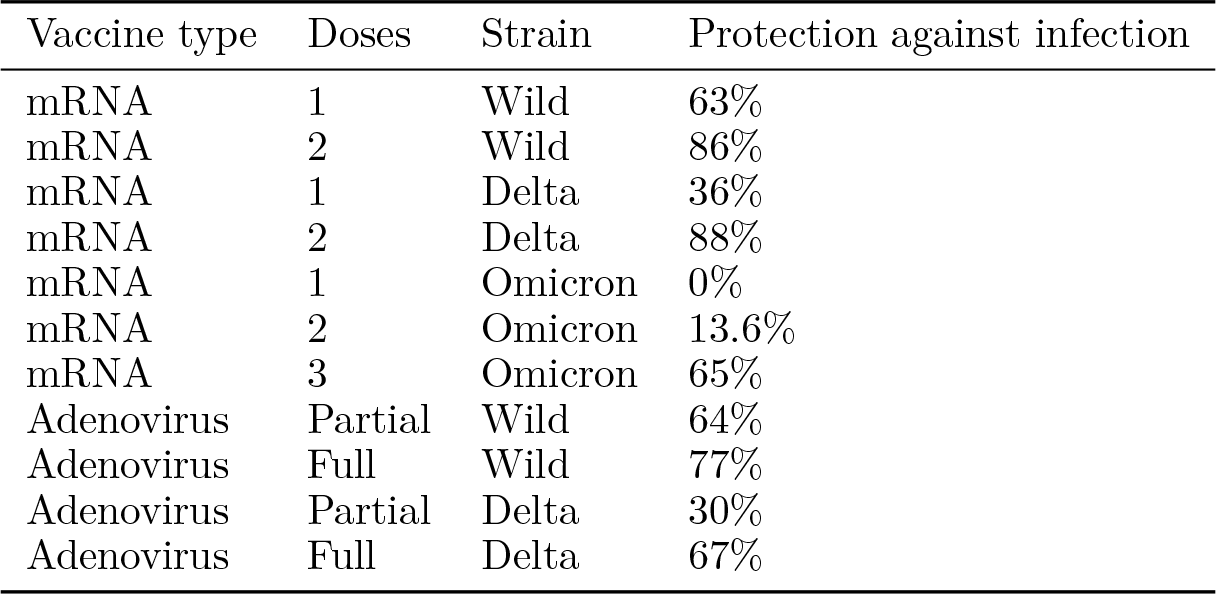
Model parameters - central estimates of vaccine efficacy

| Vaccine type | Doses | Strain | Protection against infection |
| --- | --- | --- | --- |
| mRNA | 1 | Wild | 63% |
| mRNA | 2 | Wild | 86% |
| mRNA | 1 | Delta | 36% |
| mRNA | 2 | Delta | 88% |
| mRNA | 1 | Omicron | 0% |
| mRNA | 2 | Omicron | 13.6% |
| mRNA | 3 | Omicron | 65% |
| Adenovirus | Partial | Wild | 64% |
| Adenovirus | Full | Wild | 77% |
| Adenovirus | Partial | Delta | 30% |
| Adenovirus | Full | Delta | 67% |

The shape parameters 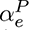, 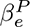 that appear in Table S1 are the shape parameters associated with a beta distribution with mean 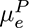 and variance 0.005. The mean 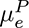 was determined by estimating the efficacy of each vaccine platform *P* against each variant (see Table S2). The mean vaccine durations were determined according to models of antibody decay and are dependent on the sampled vaccine efficacies 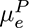 (Watson et al., 2022a). The infection fatality rates *f_min_*, *f_med_* and *f_max_* were calculated for each country according to the age-specific infection fatality rate estimates in (Brazeau et al., 2020). In these estimates, a typical high income country like the UK or the USA has an overall infection fatality rate of around 1.15% (with a 95% credible interval from 0.78% to 1.79%).

We made one change to the model used by (Watson et al., 2022a): by default, vaccination was assumed to reduce onward transmission by 50%, regardless of whether the dose was “fresh” or if the protection has waned. We modified this to the schedule shown in Table S3. These represented an average between the estimates of (Eyre et al., 2021) for effectiveness at blocking onward transmission of Alpha and Delta and the estimate of (Tan et al., 2023) for effectiveness of the vaccine at blocking onward transmission of Omicron, as the period of the simulation included both Delta and Omicron waves.

**Table S3:** Assumed relative transmissibility of infection by vaccinated individuals compared to unvaccinated

| Doses | Protection from onward transmission |
| --- | --- |
| 1 dose (not wanted) | 27% |
| 2 doses (not wanted) | 27% |
| 2 doses (waned) | 0% |
| 3 doses (not wanted) | 30% |
| 3 doses (waned stage 1) | 10% |
| 3 doses (waned stage 2) | 5% |

### D Sensitivity To Model Parameters

We explored various alternative model configurations in order to assess the sensitivity of our conclusion to modelling assumptions. Compared to a hypothetical ideal model that yields correct counterfactual assessments, the model we employ is likely to differ in a number of ways:

- It may differ structurally. We might anticipate some structural differences - for example, overdispersion in the distribution of contact rates is not captured by our model, but overdispersion in this distribution was typically found to be high (Endo et al., 2020). However, there may be other structural differences that we do not anticipate
- The assumed distributions of input parameters may differ in our model and in the hypothetical ideal

Both of these differences mean that the fitted values of *R_t_* are also likely to differ between our model and the hypothetical ideal, and hence they may yield substantially different assessments of counterfactual scenarios. If it turns out that counterfactual assessments are very sensitive to input parameter values, then we might conclude that our estimates are likely to differ substantially from the “true” counterfactual. Because of the prospect of structural differences, this is true even if we have done a very good job of estimating parameters. If our results are robust to variation of parameters within a reasonable range, then if we believe that our model is capable of yielding a good approximation to the ideal for some “reasonable” parameter choices, we should also think our counterfactual assessments are a good approximation to ideal counterfactual assessments.

We assess the impact of three different parameter estimates on our overall results. To do this, we examine the average conclusion from the model runs featuring the top and bottom deciles of each of the following parameters:

- The average infection-blocking efficacy of one and two doses of the vaccine
- The average duration for protection due to one and two doses of the vaccine
- The average duration of protection due to natural immunity

In both countries, the estimates of deaths averted showed little sensitivity to the estimated duration of natural immunity (Table S6. The estimates of deaths averted in the UK was *also* relatively insensitive to the estimated vaccine efficacy, though the estimate in the US was more sensitive to this value (it is worth noting that in the case of the US the model explored captured a wider range of variation in vaccine efficacy) - see Table S4. Note that for long-run estimates of deaths averted in the US (up to Jan 2022), higher estimates of vaccine efficacy were associated with much greater uncertainty over the number of deaths averted. This may be due to the fact that, given a more effective vaccine, waning immunity will have a larger impact on the end results.

The estimate of long-run deaths averted in the US was extremely sensitive to the estimated duration of vaccine derived immunity, with a difference of 14 days in this parameter estimate yielding long-run estimates of deaths averted that ranged from 32,255 to *negative* 11,456 (that is, 11,456 extra deaths) for a 30-day advance in the vaccination schedule. Notably, short estimates of vaccine duration were also associated with extreme decreases in the uncertainty over the number of deaths averted. Note that the short run estimates of deaths averted (up to July 2021) were robustly positive, but were also substantially more uncertain for longer estimates of the duration of vaccine protection.

Our model is already very uncertain about the effect of earlier vaccinations on the second infection wave in the US. However, the high sensitivity of this figure to the estimated duration of vaccine-derived protection offers an extra reason to be unsure that the model is providing us with an accurate assessment of the counterfactual impacts on this timescale.

We also run an identical analysis to our main analysis, except with a model fit to reported numbers of COVID-19 deaths instead of COVID-19 deaths estimated from excess mortality. The results are reported in Table S11 and Figure S6. This method yields larger estimates of deaths averted than our main method, particularly up to July 2021 where the estimates are close to the 95th percentile estimate for the main method. This is in spite of the fact that the total estimated number of deaths under this method is somewhat lower than under the excess mortality method.

**Table S4:**
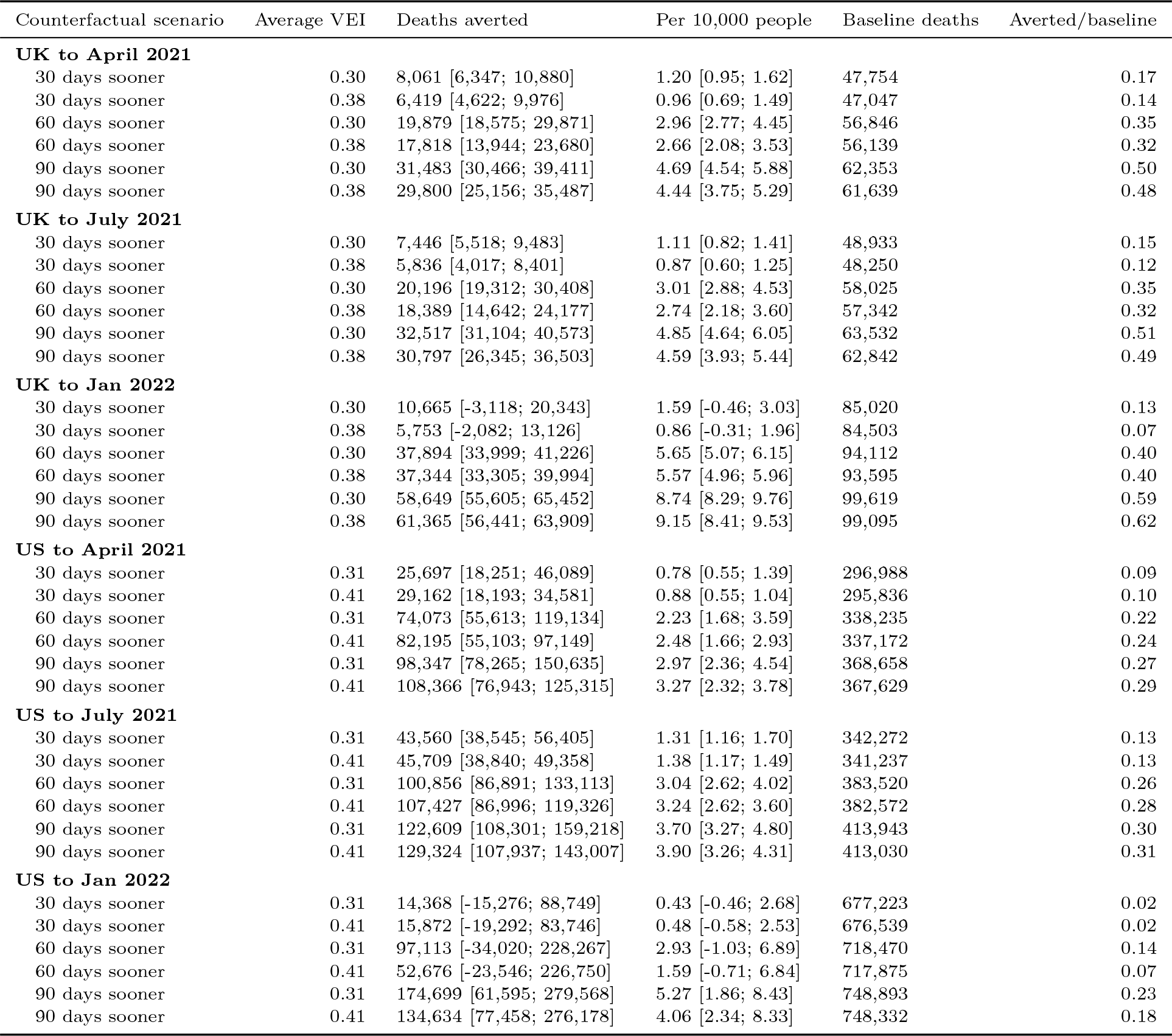
Sensitivity to average vaccine efficacy against infection (VEI)

| Counterfactual scenario | Average VEI | Deaths averted | Per 10,000 people | Baseline deaths | Averted/baseline |
| --- | --- | --- | --- | --- | --- |
| **UK to April 2021** |  |  |  |  |  |
| 30 days sooner | 0.30 | 8,061 [6,347; 10,880] | 1.20 [0.95; 1.62] | 47,754 | 0.17 |
| 30 days sooner | 0.38 | 6,419 [4,622; 9,976] | 0.96 [0.69; 1.49] | 47,047 | 0.14 |
| 60 days sooner | 0.30 | 19,879 [18,575; 29,871] | 2.96 [2.77; 4.45] | 56,846 | 0.35 |
| 60 days sooner | 0.38 | 17,818 [13,944; 23,680] | 2.66 [2.08; 3.53] | 56,139 | 0.32 |
| 90 days sooner | 0.30 | 31,483 [30,466; 39,411] | 4.69 [4.54; 5.88] | 62,353 | 0.50 |
| 90 days sooner | 0.38 | 29,800 [25,156; 35,487] | 4.44 [3.75; 5.29] | 61,639 | 0.48 |
| **UK to July 2021** |  |  |  |  |  |
| 30 days sooner | 0.30 | 7,446 [5,518; 9,483] | 1.11 [0.82; 1.41] | 48,933 | 0.15 |
| 30 days sooner | 0.38 | 5,836 [4,017; 8,401] | 0.87 [0.60; 1.25] | 48,250 | 0.12 |
| 60 days sooner | 0.30 | 20,196 [19,312; 30,408] | 3.01 [2.88; 4.53] | 58,025 | 0.35 |
| 60 days sooner | 0.38 | 18,389 [14,642; 24,177] | 2.74 [2.18; 3.60] | 57,342 | 0.32 |
| 90 days sooner | 0.30 | 32,517 [31,104; 40,573] | 4.85 [4.64; 6.05] | 63,532 | 0.51 |
| 90 days sooner | 0.38 | 30,797 [26,345; 36,503] | 4.59 [3.93; 5.44] | 62,842 | 0.49 |
| **UK to Jan 2022** |  |  |  |  |  |
| 30 days sooner | 0.30 | 10,665 [-3,118; 20,343] | 1.59 [-0.46; 3.03] | 85,020 | 0.13 |
| 30 days sooner | 0.38 | 5,753 [-2,082; 13,126] | 0.86 [-0.31; 1.96] | 84,503 | 0.07 |
| 60 days sooner | 0.30 | 37,894 [33,999; 41,226] | 5.65 [5.07; 6.15] | 94,112 | 0.40 |
| 60 days sooner | 0.38 | 37,344 [33,305; 39,994] | 5.57 [4.96; 5.96] | 93,595 | 0.40 |
| 90 days sooner | 0.30 | 58,649 [55,605; 65,452] | 8.74 [8.29; 9.76] | 99,619 | 0.59 |
| 90 days sooner | 0.38 | 61,365 [56,441; 63,909] | 9.15 [8.41; 9.53] | 99,095 | 0.62 |
| **US to April 2021** |  |  |  |  |  |
| 30 days sooner | 0.31 | 25,697 [18,251; 46,089] | 0.78 [0.55; 1.39] | 296,988 | 0.09 |
| 30 days sooner | 0.41 | 29,162 [18,193; 34,581] | 0.88 [0.55; 1.04] | 295,836 | 0.10 |
| 60 days sooner | 0.31 | 74,073 [55,613; 119,134] | 2.23 [1.68; 3.59] | 338,235 | 0.22 |
| 60 days sooner | 0.41 | 82,195 [55,103; 97,149] | 2.48 [1.66; 2.93] | 337,172 | 0.24 |
| 90 days sooner | 0.31 | 98,347 [78,265; 150,635] | 2.97 [2.36; 4.54] | 368,658 | 0.27 |
| 90 days sooner | 0.41 | 108,366 [76,943; 125,315] | 3.27 [2.32; 3.78] | 367,629 | 0.29 |
| **US to July 2021** |  |  |  |  |  |
| 30 days sooner | 0.31 | 43,560 [38,545; 56,405] | 1.31 [1.16; 1.70] | 342,272 | 0.13 |
| 30 days sooner | 0.41 | 45,709 [38,840; 49,358] | 1.38 [1.17; 1.49] | 341,237 | 0.13 |
| 60 days sooner | 0.31 | 100,856 [86,891; 133,113] | 3.04 [2.62; 4.02] | 383,520 | 0.26 |
| 60 days sooner | 0.41 | 107,427 [86,996; 119,326] | 3.24 [2.62; 3.60] | 382,572 | 0.28 |
| 90 days sooner | 0.31 | 122,609 [108,301; 159,218] | 3.70 [3.27; 4.80] | 413,943 | 0.30 |
| 90 days sooner | 0.41 | 129,324 [107,937; 143,007] | 3.90 [3.26; 4.31] | 413,030 | 0.31 |
| **US to Jan 2022** |  |  |  |  |  |
| 30 days sooner | 0.31 | 14,368 [-15,276; 88,749] | 0.43 [-0.46; 2.68] | 677,223 | 0.02 |
| 30 days sooner | 0.41 | 15,872 [-19,292; 83,746] | 0.48 [-0.58; 2.53] | 676,539 | 0.02 |
| 60 days sooner | 0.31 | 97,113 [-34,020; 228,267] | 2.93 [-1.03; 6.89] | 718,470 | 0.14 |
| 60 days sooner | 0.41 | 52,676 [-23,546; 226,750] | 1.59 [-0.71; 6.84] | 717,875 | 0.07 |
| 90 days sooner | 0.31 | 174,699 [61,595; 279,568] | 5.27 [1.86; 8.43] | 748,893 | 0.23 |
| 90 days sooner | 0.41 | 134,634 [77,458; 276,178] | 4.06 [2.34; 8.33] | 748,332 | 0.18 |

**Figure S2:**
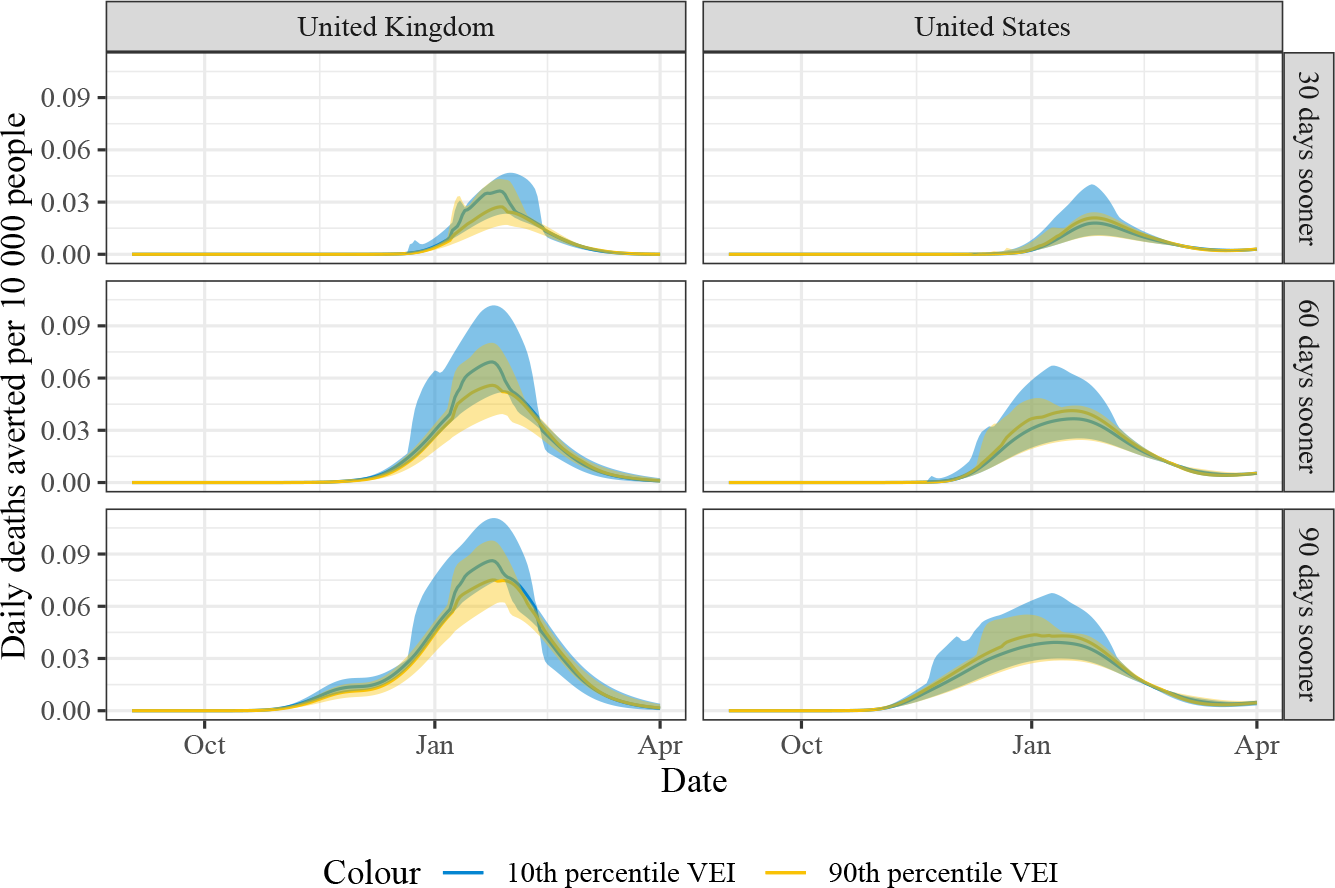
Daily averted deaths under counterfactual scenarios, with sensitivity to vaccine efficacy. Shaded area indicates 95% confidence interval.

**Table S5:** Sensitivity to duration of vaccine acquired immunity (DVI)

| Counterfactual scenario | Average DVI | Deaths averted | Per 10,000 people | Baseline deaths | Averted/baseline |
| --- | --- | --- | --- | --- | --- |
| **UK to April 2021** |  |  |  |  |  |
| 30 days sooner | 307 | 6,714 [5,833; 7,661] | 1.00 [0.87; 1.14] | 47,200 | 0.14 |
| 30 days sooner | 311 | 7,731 [5,861; 11,161] | 1.15 [0.87; 1.66] | 47,581 | 0.16 |
| 60 days sooner | 307 | 18,401 [17,170; 19,475] | 2.74 [2.56; 2.90] | 56,302 | 0.33 |
| 60 days sooner | 311 | 19,835 [16,898; 28,831] | 2.96 [2.52; 4.30] | 56,695 | 0.35 |
| 90 days sooner | 307 | 30,356 [28,784; 31,251] | 4.53 [4.29; 4.66] | 61,807 | 0.49 |
| 90 days sooner | 311 | 31,177 [28,748; 38,715] | 4.65 [4.29; 5.77] | 62,209 | 0.50 |
| **UK to July 2021** |  |  |  |  |  |
| 30 days sooner | 307 | 6,067 [5,116; 6,892] | 0.90 [0.76; 1.03] | 48,481 | 0.13 |
| 30 days sooner | 311 | 7,092 [5,130; 10,198] | 1.06 [0.76; 1.52] | 48,759 | 0.15 |
| 60 days sooner | 307 | 18,998 [17,896; 19,932] | 2.83 [2.67; 2.97] | 57,583 | 0.33 |
| 60 days sooner | 311 | 20,188 [17,492; 29,002] | 3.01 [2.61; 4.32] | 57,873 | 0.35 |
| 90 days sooner | 307 | 31,184 [29,588; 32,469] | 4.65 [4.41; 4.84] | 63,088 | 0.49 |
| 90 days sooner | 311 | 31,860 [29,786; 39,116] | 4.75 [4.44; 5.83] | 63,386 | 0.50 |
| **UK to Jan 2022** |  |  |  |  |  |
| 30 days sooner | 307 | 5,361 [-787; 6,472] | 0.80 [-0.12; 0.96] | 84,738 | 0.06 |
| 30 days sooner | 311 | 10,736 [869; 16,592] | 1.60 [0.13; 2.47] | 84,932 | 0.13 |
| 60 days sooner | 307 | 39,224 [36,623; 40,855] | 5.85 [5.46; 6.09] | 93,841 | 0.42 |
| 60 days sooner | 311 | 37,621 [33,808; 40,629] | 5.61 [5.04; 6.06] | 94,046 | 0.40 |
| 90 days sooner | 307 | 63,745 [60,012; 64,470] | 9.50 [8.95; 9.61] | 99,345 | 0.64 |
| 90 days sooner | 311 | 60,462 [54,577; 63,479] | 9.01 [8.14; 9.46] | 99,560 | 0.61 |
| **US to April 2021** |  |  |  |  |  |
| 30 days sooner | 316 | 20,583 [18,676; 33,860] | 0.62 [0.56; 1.02] | 297,055 | 0.07 |
| 30 days sooner | 319 | 27,058 [19,662; 41,157] | 0.82 [0.59; 1.24] | 296,599 | 0.09 |
| 60 days sooner | 316 | 62,213 [57,741; 96,328] | 1.88 [1.74; 2.91] | 338,265 | 0.18 |
| 60 days sooner | 319 | 77,933 [59,667; 111,234] | 2.35 [1.80; 3.36] | 337,870 | 0.23 |
| 90 days sooner | 316 | 86,836 [81,229; 123,798] | 2.62 [2.45; 3.73] | 368,675 | 0.24 |
| 90 days sooner | 319 | 103,449 [83,147; 143,490] | 3.12 [2.51; 4.33] | 368,297 | 0.28 |
| **US to July 2021** |  |  |  |  |  |
| 30 days sooner | 316 | 40,960 [39,705; 48,919] | 1.24 [1.20; 1.48] | 342,380 | 0.12 |
| 30 days sooner | 319 | 45,377 [40,193; 51,652] | 1.37 [1.21; 1.56] | 341,852 | 0.13 |
| 60 days sooner | 316 | 93,289 [89,688; 118,305] | 2.81 [2.71; 3.57] | 383,589 | 0.24 |
| 60 days sooner | 319 | 105,808 [91,011; 125,079] | 3.19 [2.75; 3.77] | 383,123 | 0.28 |
| 90 days sooner | 316 | 116,022 [110,928; 140,186] | 3.50 [3.35; 4.23] | 414,000 | 0.28 |
| 90 days sooner | 319 | 127,677 [113,003; 152,312] | 3.85 [3.41; 4.59] | 413,550 | 0.31 |
| **US to Jan 2022** |  |  |  |  |  |
| 30 days sooner | 316 | 20,707 [-15,079; 79,958] | 0.62 [-0.45; 2.41] | 677,770 | 0.03 |
| 30 days sooner | 319 | 15,258 [-16,320; 56,716] | 0.46 [-0.49; 1.71] | 676,626 | 0.02 |
| 60 days sooner | 316 | 104,249 [-19,159; 221,533] | 3.14 [-0.58; 6.68] | 718,979 | 0.14 |
| 60 days sooner | 319 | 78,202 [-19,587; 182,171] | 2.36 [-0.59; 5.50] | 717,897 | 0.11 |
| 90 days sooner | 316 | 181,420 [79,392; 275,209] | 5.47 [2.39; 8.30] | 749,389 | 0.24 |
| 90 days sooner | 319 | 162,718 [82,397; 243,528] | 4.91 [2.49; 7.35] | 748,323 | 0.22 |

**Figure S3:**
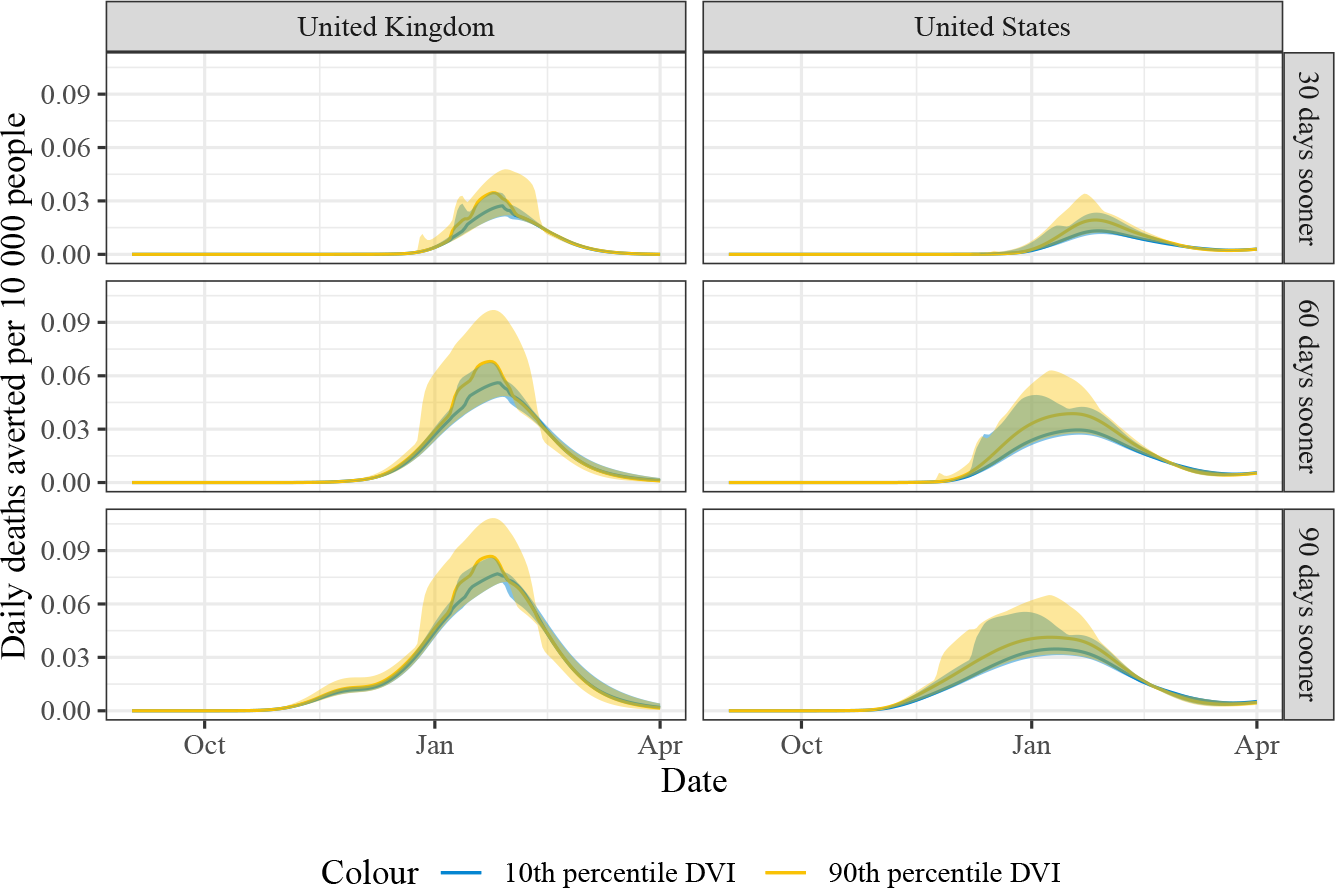
Daily averted deaths under counterfactual scenarios, with sensitivity to vaccine duration. Shaded area indicates 95% confidence interval.

**Table S6:**
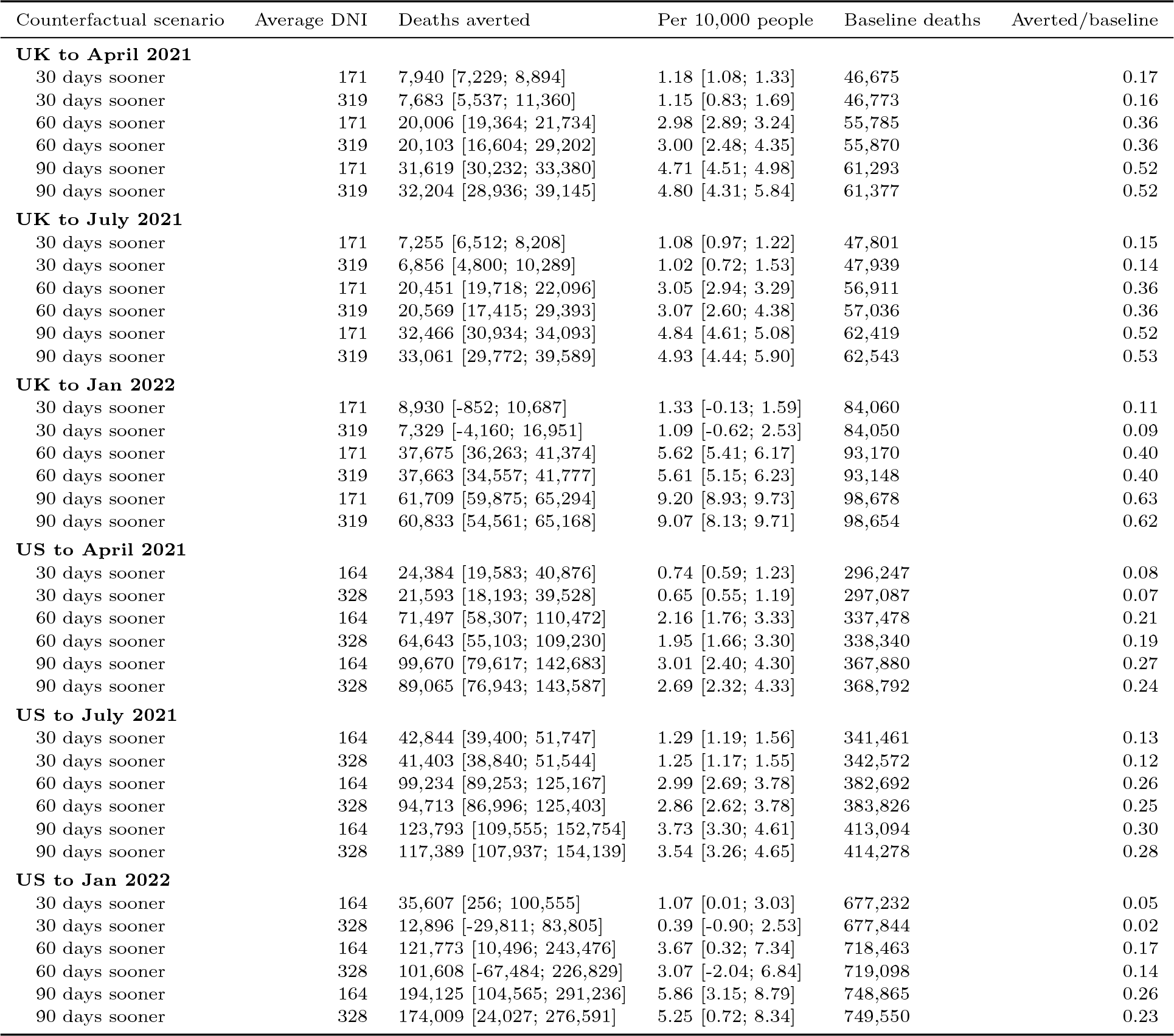
Sensitivity to duration of naturally acquired immunity (DNI)

**Figure S4:**
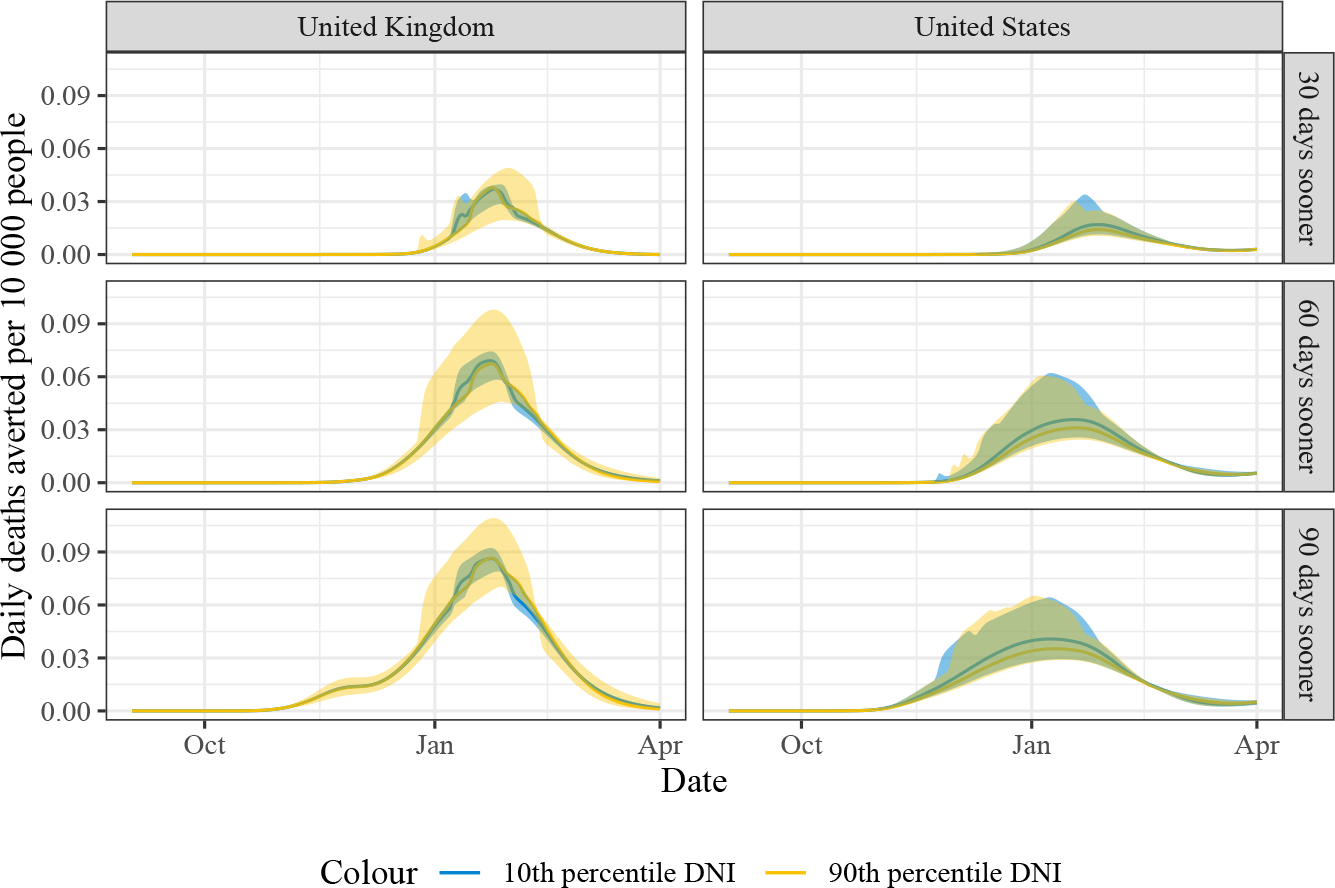
Daily averted deaths under counterfactual scenarios, with sensitivity to natural immunity duration. Shaded area indicates 95% confidence interval.

**Figure S5:**
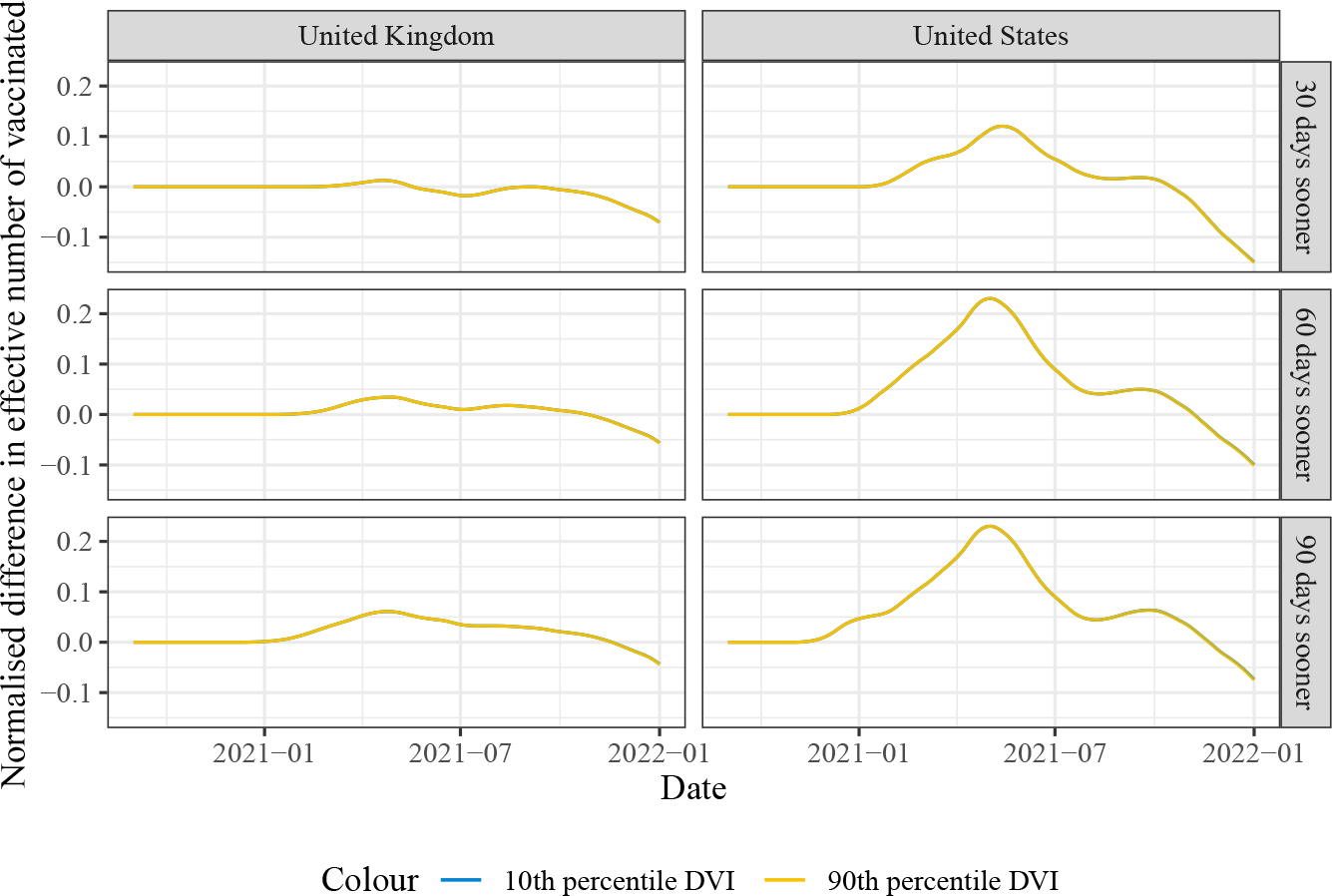
Effective vaccinated, sensitivity to vaccine duration

### E Supplementary Figures and Tables

**Figure S6:**
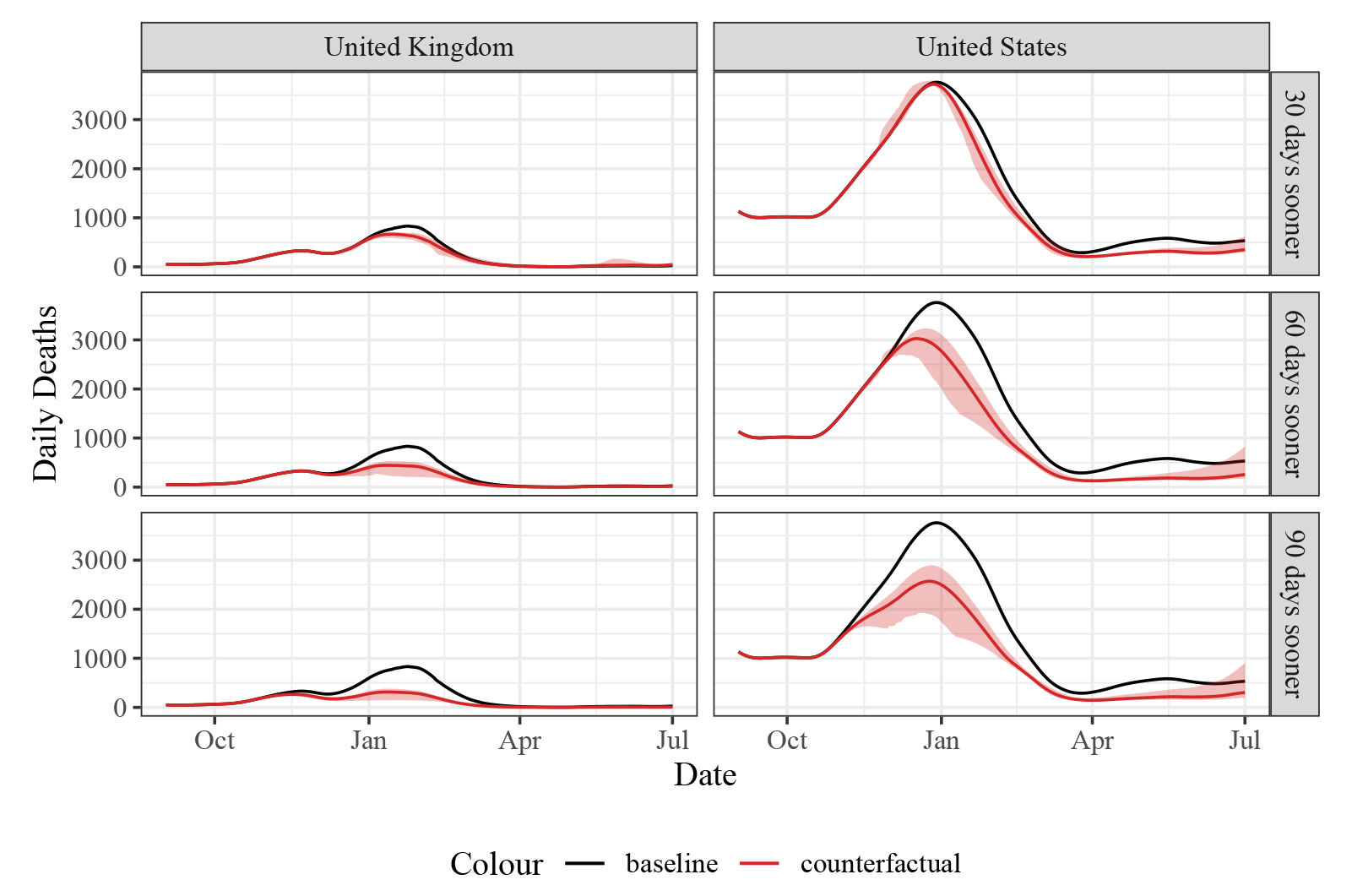
Reported deaths for each counerfactual scenario compared to baseline reported deaths. Shaded area indicates 95% confidence interval.

**Table S7:** Total averted deaths under counterfactual scenarios, based on reported deaths. 95% intervals are calculated over 100 simulation runs which vary vaccine parameters and duration of natural immunity (see Table S1 for details). This table provides additional results for Table 2.

| Counterfactual scenario | Deaths averted | Per 10,000 people | Baseline deaths | Averted/baseline |
| --- | --- | --- | --- | --- |
| **United Kingdom to April 2021** |  |  |  |  |
| 30 days sooner | 10,728 [6,627; 14,991] | 1.60 [0.99; 2.23] | 69,808 | 0.15 |
| 60 days sooner | 29,922 [21,040; 43,472] | 4.46 [3.14; 6.48] | 84,124 | 0.36 |
| 90 days sooner | 46,665 [37,966; 67,151] | 6.96 [5.66; 10.01] | 92,216 | 0.51 |
| **United Kingdom to July 2021** |  |  |  |  |
| 30 days sooner | 9,952 [6,026; 13,949] | 1.48 [0.90; 2.08] | 71,862 | 0.14 |
| 60 days sooner | 30,516 [21,967; 43,452] | 4.55 [3.27; 6.48] | 86,178 | 0.35 |
| 90 days sooner | 47,953 [39,529; 67,086] | 7.15 [5.89; 10.00] | 94,270 | 0.51 |
| **United Kingdom to Jan 2022** |  |  |  |  |
| 30 days sooner | 8,019 [-2,350; 17,181] | 1.20 [-0.35; 2.56] | 95,018 | 0.08 |
| 60 days sooner | 41,389 [35,940; 49,167] | 6.17 [5.36; 7.33] | 109,334 | 0.38 |
| 90 days sooner | 65,485 [60,730; 77,178] | 9.76 [9.05; 11.51] | 117,427 | 0.56 |
| **United States to April 2021** |  |  |  |  |
| 30 days sooner | 35,207 [25,342; 63,091] | 1.06 [0.76; 1.90] | 310,390 | 0.11 |
| 60 days sooner | 87,601 [67,671; 143,053] | 2.64 [2.04; 4.32] | 341,910 | 0.26 |
| 90 days sooner | 107,134 [86,198; 167,087] | 3.23 [2.60; 5.04] | 364,108 | 0.29 |
| **United States to July 2021** |  |  |  |  |
| 30 days sooner | 53,342 [45,493; 73,576] | 1.61 [1.37; 2.22] | 358,199 | 0.15 |
| 60 days sooner | 115,895 [99,725; 152,977] | 3.50 [3.01; 4.61] | 389,719 | 0.30 |
| 90 days sooner | 132,919 [117,226; 173,179] | 4.01 [3.54; 5.22] | 411,917 | 0.32 |
| **United States to Jan 2022** |  |  |  |  |
| 30 days sooner | 45,196 [3,320; 101,695] | 1.36 [0.10; 3.07] | 583,198 | 0.08 |
| 60 days sooner | 120,655 [-32,178; 221,714] | 3.64 [-0.97; 6.69] | 614,719 | 0.20 |
| 90 days sooner | 175,230 [56,961; 255,219] | 5.29 [1.72; 7.70] | 636,916 | 0.28 |

**Figure S7:**
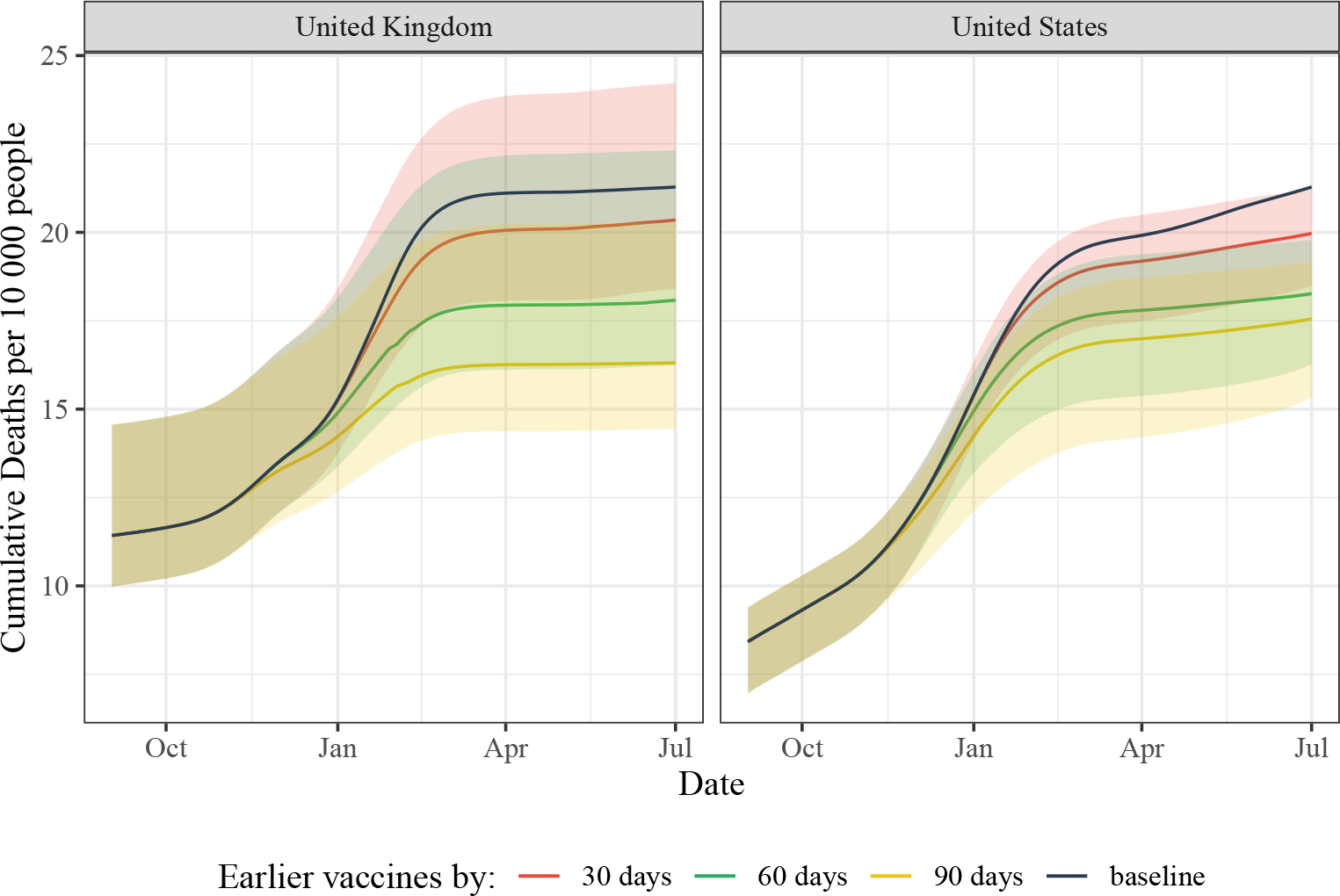
Cumulative deaths under baseline and counterfactual scenarios. Shaded area indicates 95% confidence interval

**Table S8:** How would outcomes change if vaccination demand were 10% lower

| Counterfactual scenario | Deaths averted | Per 10,000 people | Baseline deaths | Averted/baseline |
| --- | --- | --- | --- | --- |
| **United Kingdom to April 2021** |  |  |  |  |
| 30 days sooner | 9,208 [5,600; 12,392] | 1.37 [0.83; 1.85] | 69,808 | 0.13 |
| 60 days sooner | 27,691 [19,208; 39,888] | 4.13 [2.86; 5.95] | 84,124 | 0.33 |
| 90 days sooner | 44,061 [35,169; 65,092] | 6.57 [5.24; 9.70] | 92,216 | 0.48 |
| **United Kingdom to July 2021** |  |  |  |  |
| 30 days sooner | 7,596 [4,228; 10,548] | 1.13 [0.63; 1.57] | 71,862 | 0.11 |
| 60 days sooner | 27,735 [19,660; 39,240] | 4.13 [2.93; 5.85] | 86,178 | 0.32 |
| 90 days sooner | 44,886 [36,445; 64,154] | 6.69 [5.43; 9.56] | 94,270 | 0.48 |
| **United Kingdom to Jan 2022** |  |  |  |  |
| 30 days sooner | -14,878 [-76,737; 7,384] | -2.22 [-11.44; 1.10] | 95,018 | -0.16 |
| 60 days sooner | 9,691 [-14,387; 29,963] | 1.44 [-2.14; 4.47] | 109,334 | 0.09 |
| 90 days sooner | 39,790 [31,759; 56,986] | 5.93 [4.73; 8.50] | 117,427 | 0.34 |
| **United States to April 2021** |  |  |  |  |
| 30 days sooner | 30,365 [21,689; 55,905] | 0.92 [0.65; 1.69] | 310,390 | 0.10 |
| 60 days sooner | 80,279 [61,435; 133,348] | 2.42 [1.85; 4.02] | 341,910 | 0.23 |
| 90 days sooner | 98,186 [79,051; 155,592] | 2.96 [2.38; 4.69] | 364,108 | 0.27 |
| **United States to July 2021** |  |  |  |  |
| 30 days sooner | 40,202 [33,996; 58,510] | 1.21 [1.03; 1.77] | 358,199 | 0.11 |
| 60 days sooner | 101,539 [87,394; 134,921] | 3.06 [2.64; 4.07] | 389,719 | 0.26 |
| 90 days sooner | 115,630 [104,019; 153,078] | 3.49 [3.14; 4.62] | 411,917 | 0.28 |
| **United States to Jan 2022** |  |  |  |  |
| 30 days sooner | -49,504 [-90,132; 32,157] | -1.49 [-2.72; 0.97] | 583,198 | -0.08 |
| 60 days sooner | 23,485 [-71,246; 90,271] | 0.71 [-2.15; 2.72] | 614,719 | 0.04 |
| 90 days sooner | 80,385 [6,176; 143,145] | 2.42 [0.19; 4.32] | 636,916 | 0.13 |

**Table S9:** How would outcomes change if vaccination demand were 5% lower?

| Counterfactual scenario | Deaths averted | Per 10,000 people | Baseline deaths | Averted/baseline |
| --- | --- | --- | --- | --- |
| **United Kingdom to April 2021** |  |  |  |  |
| 30 days sooner | 9,977 [6,147; 13,710] | 1.49 [0.92; 2.04] | 69,808 | 0.14 |
| 60 days sooner | 28,797 [20,136; 41,753] | 4.29 [3.00; 6.22] | 84,124 | 0.34 |
| 90 days sooner | 45,385 [36,575; 66,146] | 6.77 [5.45; 9.86] | 92,216 | 0.49 |
| **United Kingdom to July 2021** |  |  |  |  |
| 30 days sooner | 8,802 [5,190; 12,319] | 1.31 [0.77; 1.84] | 71,862 | 0.12 |
| 60 days sooner | 29,150 [20,849; 41,441] | 4.35 [3.11; 6.18] | 86,178 | 0.34 |
| 90 days sooner | 46,465 [38,005; 65,640] | 6.93 [5.67; 9.79] | 94,270 | 0.49 |
| **United Kingdom to Jan 2022** |  |  |  |  |
| 30 days sooner | -4,209 [-36,876; 11,673] | -0.63 [-5.50; 1.74] | 95,018 | -0.04 |
| 60 days sooner | 26,578 [20,035; 38,228] | 3.96 [2.99; 5.70] | 109,334 | 0.24 |
| 90 days sooner | 56,491 [48,152; 68,647] | 8.42 [7.18; 10.23] | 117,427 | 0.48 |
| **United States to April 2021** |  |  |  |  |
| 30 days sooner | 32,814 [23,533; 59,598] | 0.99 [0.71; 1.80] | 310,390 | 0.11 |
| 60 days sooner | 84,004 [64,622; 138,347] | 2.53 [1.95; 4.17] | 341,910 | 0.25 |
| 90 days sooner | 102,734 [82,735; 161,675] | 3.10 [2.50; 4.88] | 364,108 | 0.28 |
| **United States to July 2021** |  |  |  |  |
| 30 days sooner | 47,149 [40,040; 66,332] | 1.42 [1.21; 2.00] | 358,199 | 0.13 |
| 60 days sooner | 108,980 [93,903; 144,340] | 3.29 [2.83; 4.35] | 389,719 | 0.28 |
| 90 days sooner | 125,010 [110,971; 163,692] | 3.77 [3.35; 4.94] | 411,917 | 0.30 |
| **United States to Jan 2022** |  |  |  |  |
| 30 days sooner | 2,578 [-33,216; 46,710] | 0.08 [-1.00; 1.41] | 583,198 | 0.00 |
| 60 days sooner | 74,985 [-50,099; 147,926] | 2.26 [-1.51; 4.46] | 614,719 | 0.12 |
| 90 days sooner | 128,759 [38,392; 191,873] | 3.88 [1.16; 5.79] | 636,916 | 0.20 |

**Table S10:** How would US outcomes change if boosters were adopted at twice the actual rate?

| Counterfactual scenario | Deaths averted | Per 10,000 people | Baseline deaths | Averted/baseline |
| --- | --- | --- | --- | --- |
| **United Kingdom to April 2021** |  |  |  |  |
| 30 days sooner | 6,923 [5,482; 10,845] | 1.03 [0.82; 1.62] | 47,105 | 0.15 |
| 60 days sooner | 19,055 [16,178; 29,735] | 2.84 [2.41; 4.43] | 56,206 | 0.34 |
| 90 days sooner | 31,079 [27,164; 39,334] | 4.63 [4.05; 5.86] | 61,709 | 0.50 |
| **United Kingdom to July 2021** |  |  |  |  |
| 30 days sooner | 6,256 [4,739; 9,296] | 0.93 [0.71; 1.39] | 48,329 | 0.13 |
| 60 days sooner | 19,513 [16,767; 29,889] | 2.91 [2.50; 4.46] | 57,430 | 0.34 |
| 90 days sooner | 32,028 [27,911; 39,828] | 4.77 [4.16; 5.94] | 62,933 | 0.51 |
| **United Kingdom to Jan 2022** |  |  |  |  |
| 30 days sooner | 6,240 [-4,129; 19,744] | 0.93 [-0.62; 2.94] | 84,531 | 0.07 |
| 60 days sooner | 38,111 [33,509; 41,848] | 5.68 [5.00; 6.24] | 93,631 | 0.41 |
| 90 days sooner | 62,520 [54,053; 66,192] | 9.32 [8.06; 9.87] | 99,134 | 0.63 |
| **United States to April 2021** |  |  |  |  |
| 30 days sooner | 24,552 [17,774; 40,322] | 0.74 [0.54; 1.22] | 296,895 | 0.08 |
| 60 days sooner | 72,072 [54,156; 111,904] | 2.17 [1.63; 3.38] | 338,137 | 0.21 |
| 90 days sooner | 97,464 [76,963; 149,298] | 2.94 [2.32; 4.50] | 368,555 | 0.26 |
| **United States to July 2021** |  |  |  |  |
| 30 days sooner | 43,602 [38,591; 52,063] | 1.32 [1.16; 1.57] | 342,236 | 0.13 |
| 60 days sooner | 100,332 [86,295; 126,194] | 3.03 [2.60; 3.81] | 383,478 | 0.26 |
| 90 days sooner | 124,047 [107,995; 157,820] | 3.74 [3.26; 4.76] | 413,895 | 0.30 |
| **United States to Jan 2022** |  |  |  |  |
| 30 days sooner | 51,045 [-36,144; 124,584] | 1.54 [-1.09; 3.76] | 647,587 | 0.08 |
| 60 days sooner | 160,448 [-65,642; 272,969] | 4.84 [-1.98; 8.23] | 688,829 | 0.23 |
| 90 days sooner | 254,410 [96,230; 327,270] | 7.67 [2.90; 9.87] | 719,246 | 0.35 |

**Table S11:** Averted deaths calculated on the basis of a model fit to excess deaths

| Counterfactual scenario | Deaths averted | Per 10,000 people | Baseline deaths | Averted/baseline |
| --- | --- | --- | --- | --- |
| **United Kingdom to April 2021** |  |  |  |  |
| 30 days sooner | 6,923 [5,482; 10,845] | 1.03 [0.82; 1.62] | 47,105 | 0.15 |
| 60 days sooner | 19,055 [16,178; 29,735] | 2.84 [2.41; 4.43] | 56,206 | 0.34 |
| 90 days sooner | 31,079 [27,164; 39,334] | 4.63 [4.05; 5.86] | 61,709 | 0.50 |
| **United Kingdom to July 2021** |  |  |  |  |
| 30 days sooner | 6,256 [4,739; 9,296] | 0.93 [0.71; 1.39] | 48,329 | 0.13 |
| 60 days sooner | 19,513 [16,767; 29,889] | 2.91 [2.50; 4.46] | 57,430 | 0.34 |
| 90 days sooner | 32,028 [27,911; 39,828] | 4.77 [4.16; 5.94] | 62,933 | 0.51 |
| **United Kingdom to Jan 2022** |  |  |  |  |
| 30 days sooner | 6,240 [-4,129; 19,744] | 0.93 [-0.62; 2.94] | 84,531 | 0.07 |
| 60 days sooner | 38,111 [33,509; 41,848] | 5.68 [5.00; 6.24] | 93,631 | 0.41 |
| 90 days sooner | 62,520 [54,053; 66,192] | 9.32 [8.06; 9.87] | 99,134 | 0.63 |
| **United States to April 2021** |  |  |  |  |
| 30 days sooner | 24,555 [17,788; 40,319] | 0.74 [0.54; 1.22] | 296,896 | 0.08 |
| 60 days sooner | 72,049 [54,157; 111,909] | 2.17 [1.63; 3.38] | 338,138 | 0.21 |
| 90 days sooner | 97,463 [76,943; 149,291] | 2.94 [2.32; 4.50] | 368,555 | 0.26 |
| **United States to July 2021** |  |  |  |  |
| 30 days sooner | 43,592 [38,604; 52,077] | 1.31 [1.16; 1.57] | 342,242 | 0.13 |
| 60 days sooner | 100,312 [86,296; 126,203] | 3.03 [2.60; 3.81] | 383,484 | 0.26 |
| 90 days sooner | 124,026 [107,939; 157,734] | 3.74 [3.26; 4.76] | 413,901 | 0.30 |
| **United States to Jan 2022** |  |  |  |  |
| 30 days sooner | 25,971 [-42,451; 98,451] | 0.78 [-1.28; 2.97] | 677,535 | 0.04 |
| 60 days sooner | 97,944 [-92,089; 245,497] | 2.95 [-2.78; 7.41] | 718,776 | 0.14 |
| 90 days sooner | 175,542 [12,829; 293,716] | 5.30 [0.39; 8.86] | 749,194 | 0.23 |

## Footnotes

1 In the US, in situations in which human clinical trial testing may not be feasible or unethical, animal trials may be used in place of clinical trials (referred to as the “animal rule”) or patients who have sufficient need for a possible therapy (such as terminally ill patients) so as to forgo possible safety or efficacy concerns may receive experimental drugs (referred to as “compassionate use”) (FDA, 2023, Rizk et al. (2021)). However, the obvious deficit of these approaches is that they are not based on much weaker efficacy and safety data than a EUA pathway.

2 In HCTs a small group of volunteers are intentionally infected with a disease in order to study the efficacy of potential treatments or vaccines. While both ethical and technological barriers to adoption of HCTs are considerable (as we will discuss in more detail later), they have the potential to significantly accelerate the efficacy studies, at least in theory.

3 We discuss other estimates of benefits of speed at the end of the paper.

4 The model we use could also estimate reductions in infections. Similarly, we could estimate reductions in hospitalizations or severe cases. However, for simplicity we focus only on mortality, since the relative reductions in all outcomes can be thought of as roughly comparable, and averting deaths is typically the main variable that is of interest to public health decision makers.

5 All descriptions and modelling use data from the dataset introduced by Dong et al. (2020), which was last updated on 2023-03-10. These numbers of confirmed cases are typically a bit higher than some of the revised estimates currently available online. Additionally, for alternative modelling specification we use excess mortality data from Solstad (2023), which we will discuss below. Unless otherwise stated, by COVID-19 deaths we mean confirmed deaths, not excess deaths. Due to data gaps, for vaccinations in the UK we assumed linear increase in vaccinations from 8 December 2020 (first vaccination in the UK) and 11 January 2021 (first day from which we had daily data), matching the total number of doses distributed in that period.

6 This type of estimate crucially depends on the assumptions of individual and policy responses, since prolonging a pandemic could manifest not just as additional deaths but also in continued lockdowns or people choosing to avoid contacts. However, it highlights the benefits of vaccinations are huge.

7 This also means timing of epidemic peaks (in relation to vaccination start and in relation to each other) is an important factor in calculating health burdens and possible mortality reductions from vaccination.

8 However, as we will discuss in more detail below, the number of both modeled (status quo) and counterfactual (no vaccines) deaths in our modified model is similar to the original Watson et al. paper: where the Watson paper estimates a baseline of 140 000 deaths in the UK and 800 000 deaths for the US on Dec 1 2021, our model estimates 164 000 UK deaths and 762 000 US deaths on the same date. Note that in both cases our model’s estimates are closer to published figures at the time of publication. Under the no vaccine counterfactual, the model from the Watson paper estimates 2.7 million deaths in the US compared to 2.5 million in ours, and 550 000 deaths in the UK compared to 510 000 in ours.

9 The division into typical three phases of clinical research is in this case a blurry one, since to expedite testing drug developers “merged” phases into single studies. What’s relevant to us here, however, is pinpointing the moment in time when developers would consider generating efficacy and effectiveness data for their candidate vaccines. We (roughly) settle on May 2020.

10 There are many arguments for why safety testing could be completed sooner or later than September 2020. On one hand, large-scale vaccine safety testing did not necessitate completion of phase 1/2 studies and it could have feasibly begun around April or even earlier. On the other hand, the three-month timeline is very ambitious and would have required several improvements to the already efficiently compressed trial timelines.

11 The statistic for development of a challenge agent sometimes provided by experts is of a six-month wait, but seemingly this figure is grounded less in an up-to-date feasibility assessment and more in generalizing from historical experience. For a discussion on the challenges relating to the development of challenge agents see Williams et al. (2023).

12 COVID HCTs, from the viewpoint of feasibility, risks, and value of information (on efficacy) provided have been extensively discussed elsewhere. Natural exposure trials were discussed by Eyal and Lipsitch (2021).

13 This is a conservative assumption, since production capacity was partially determined by the predicted date of regulatory approval. In other words, it is reasonable to believe that earlier approvals would have resulted in more doses being produced in 2020. However, given complexity of the vaccine supply chains early in the pandemic this may be a realistic assumption. See for example Bown and Bollyky (2022) and Kominers and Tabarrok (2022).

14 For the US we assumed that doses used were mRNA. For the UK we assumed that 50% of initial vaccinations (first two doses) were AstraZeneca vaccines and 50% were mRNA. For booster doses in both cases we assumed that mRNA vaccines were used. To do this, we use the average of vaccine efficacy parameters for AstraZeneca vaccines and mRNA. While there are no exact statistics of vaccine use in the UK broken down by manufacturer, the government press release a year on from the start of vaccinations supports this assumption (UKDH, 2022).

15 Examples of which include dynamic contact rates (e.g. over weekends and festive periods), non-homogeneous mixing (e.g. children mixing more often with other children than with the elderly), and overdispersion in contact rates (related to superspreading), none of which are reflected by the model we use here.

16 In our main period of interest, up to 1 July 2021, our results suggest that for the UK accelerating availability of vaccines would have averted between 6,300 (assuming 30-day acceleration; lower 95% bound is 4,700) and 32,000 (90-day acceleration; upper 95% bound is 40,000) deaths. This translates to 0.93-4.8 deaths averted per 10,000. In the US, corresponding numbers of deaths averted are 44,000 (lower 95% bound of 39,000) and 120,000 (upper 95% bound of 160,000) for 30-day and 90-day acceleration respectively. This gives a range of 1.3-3.7 deaths averted per 10,000.

17 In the period between 1 January and 1 July 2021 there were 61,000 confirmed COVID deaths in the UK and 246,000 in the US (Mathieu et al., 2021)).

18 The assumption of 33% chance of success (i.e. probability of regulatory approval, conditional on positive results of Phase 1/2 trials—in case of COVID-19 around May 2020) may be a slightly conservative assumption for COVID, since the vaccine developers were optimistic of approval by the time Phase 2 trials started. However, it is higher than some of the historic probabilities of clinical trial success in vaccine development (Gouglas et al., 2018).

19 See DBEI (2021). However, the cost of the HCT itself was likely closer to about £10 million (Magazine, 2020). The purpose of this human challenge trial was to conduct a characterization study rather than a vaccine study. Its primary objective was to determine an optimal inoculum dose, while the secondary objective involved evaluating the kinetics of the virus and symptoms during the infection process (Killingley et al., 2022). As an illustrative example, the rotavirus Rotarix vaccine Phase 3 trial costs are estimated to have cost between $136 million and $204 million (Light et al., 2009).

20 The World Health Organisation published eight criteria for the ethical use of SARS-CoV-2 challenge studies, which focused on scientific and ethical assessments, consultation and coordination, selection criteria, and review and consent (see Jamrozik et al. (2021))

21 This is an estimate based on Pfizer producing 2.5 times more than Moderna in 2020 but getting faster approval and running at full speed longer.

